# Large-scale trans-ethnic replication and discovery of genetic associations for rare diseases with self-reported medical data

**DOI:** 10.1101/2021.06.09.21258643

**Authors:** Suyash S. Shringarpure, Wei Wang, Yunxuan Jiang, Alison Acevedo, Devika Dhamija, Briana Cameron, Adrian Jubb, Peng Yue, The 23andMe Research Team, Lea Sarov-Blat, Robert Gentleman, Adam Auton

## Abstract

A key challenge in the study of rare disease genetics is assembling large case cohorts for well-powered studies. We demonstrate the use of self-reported diagnosis data to study rare diseases at scale. We performed genome-wide association studies (GWAS) for 33 rare diseases using self-reported diagnosis phenotypes and re-discovered 29 known associations to validate our approach. In addition, we performed the first GWAS for Duane retraction syndrome, vestibular schwannoma and spontaneous pneumothorax, and report novel genome-wide significant associations for these diseases. We replicated these novel associations in non-European populations within the 23andMe, Inc. cohort as well as in the UK Biobank cohort. We also show that mixed model analyses including all ethnicities and related samples increase the power for finding associations in rare diseases. Our results, based on analysis of 19,084 rare disease cases for 33 diseases from 7 populations, show that large-scale online collection of self-reported data is a viable method for discovery and replication of genetic associations for rare diseases. This approach, which is complementary to sequencing-based approaches, will enable the discovery of more novel genetic associations for increasingly rare diseases across multiple ancestries and shed more light on the genetic architecture of rare diseases.

## Introduction

Rare diseases are conditions that affect fewer than 200,000 people in the United States or no more than 1 of every 2,000 people in Europe, with other definitions based on varying prevalence thresholds across the world^1^. Nearly 7000 rare diseases are known, and though each disease affects a small number of people, the total population prevalence of rare diseases is estimated to be 3.5-5.9%^2^. Many rare diseases have pediatric onset, are chronic, and few are curable. Therefore, rare diseases are emerging as a public health priority^3^.

The study of the genetics of rare diseases is challenging due to their low prevalence and the difficulty in creating large case cohorts for well-powered studies. Though more than 70% of rare diseases have genetic origins, only about half of known rare diseases have at least one putatively associated gene^2^. For a few rare diseases, large cohorts have been assembled -- for example, the Genetic Modifiers of Huntington’s Disease (GeM-HD) Consortium^4^ for Huntington’s disease with 4,802 cases. For other diseases, studies have meta-analyzed multiple small cohorts^5–7^. In a recent study, whole-genome sequencing data from 9,802 rare disease patients in a national health system was used to study 15 rare disease domains^8^. For the majority of rare diseases, associated genes have been discovered by sequencing whole genomes or exomes from a small number of cases or case families, followed by automated or manual variant prioritization^9,10^. Since up to 6% of the world’s population is estimated to have a rare disease, assembling and sequencing large case populations to study many rare diseases simultaneously is a challenging problem. Complementary approaches based on large-scale data collection through self-report and scalable analyses through genome-wide association studies (GWAS) could provide additional insights into the genetic architecture of rare diseases.

Here we demonstrate the use of large-scale self-reported rare disease data, combined with genetic data collected through the 23andMe direct-to-consumer (DTC) platform, to study rare diseases at a large scale and identify genetic associations for rare diseases through GWAS. Using web-based questionnaires, we gathered self-reported data on rare diseases from a cohort of over 1.6 million genotyped research-consented individuals. Through simulations, we showed that GWAS for rare monogenic diseases are well-powered to find associations near causal genes. We ran GWAS on 33 rare disease phenotypes in individuals of European ancestry for which we had sufficient sample size. We reidentified 29 known associations at a genome-wide significance level (p-value < 5e-8) with a diverse range of minor allele frequencies (minimum MAF=0.0001, maximum MAF=0.4873) and risk allele effect sizes (minimum OR=1.2392, maximum OR=273.149). The rare diseases in which we replicate genetic associations have a variety of architectures, from monogenic to polygenic. We also replicated some of the significant associations in individuals of African-American, Latino, and East Asian ancestries. We identified novel genome-wide significant associations for orofacial clefts, vestibular schwannoma, Duane retraction syndrome and spontaneous pneumothorax, which we replicated in the UK Biobank and an independent subset of the 23andMe cohort. In addition, we show that power to find these associations can be increased by mixed-model analysis of data across all ancestries.

## Results

### Validation of the approach

To validate our approach, we examined the power of GWAS on self-reported data to replicate known associations. For this, we used simulations of monogenic rare diseases using genotyped SNPs as causal variants. We also performed GWAS on rare diseases with known causal genes to study whether known associations could be reidentified. Lastly, where GWAS on clinical rare disease cohorts were available in the GWAS catalog, we compared effect sizes between matched GWAS hits in our GWAS on self-reported data and the clinical GWAS.

#### Power of GWAS to find associations in simulated monogenic rare diseases

We examined the power of GWAS to find associations for monogenic diseases by simulating rare monogenic diseases with different genetic architectures in a cohort of 4,957,230 research-consented individuals of European ancestry (Methods and Supplementary Notes, Section 1). We found that for dominant rare diseases, GWAS was well-powered for diseases with prevalence more than 1 in 50,000 (Figure 1(a)), with GWAS power decreasing as the number of causal variants in the gene increased. GWAS power was larger when causal variants had incomplete penetrance, likely due to the larger MAF of causal variants in scenarios with incomplete penetrance and therefore an improved ability for causal variants to be tagged by variants included in the GWAS. Also, the GWAS lead SNPs from genome-wide significant hits were more common in frequency than the selected causal variants (Supplementary Figure 1(a)), indicating that the GWAS lead SNPs were not the causal variants themselves, but more common SNPs linked to the causal variants. For recessive diseases, we find that the GWAS has nearly 100% power to find an association near the causal gene in all disease architectures we simulated (Figure 1(b)). Similar to the results from simulations of dominant diseases, we find that GWAS lead SNPs are more common than selected causal variants in simulation (Supplementary Figure 1(b)).

**Figure 1:**
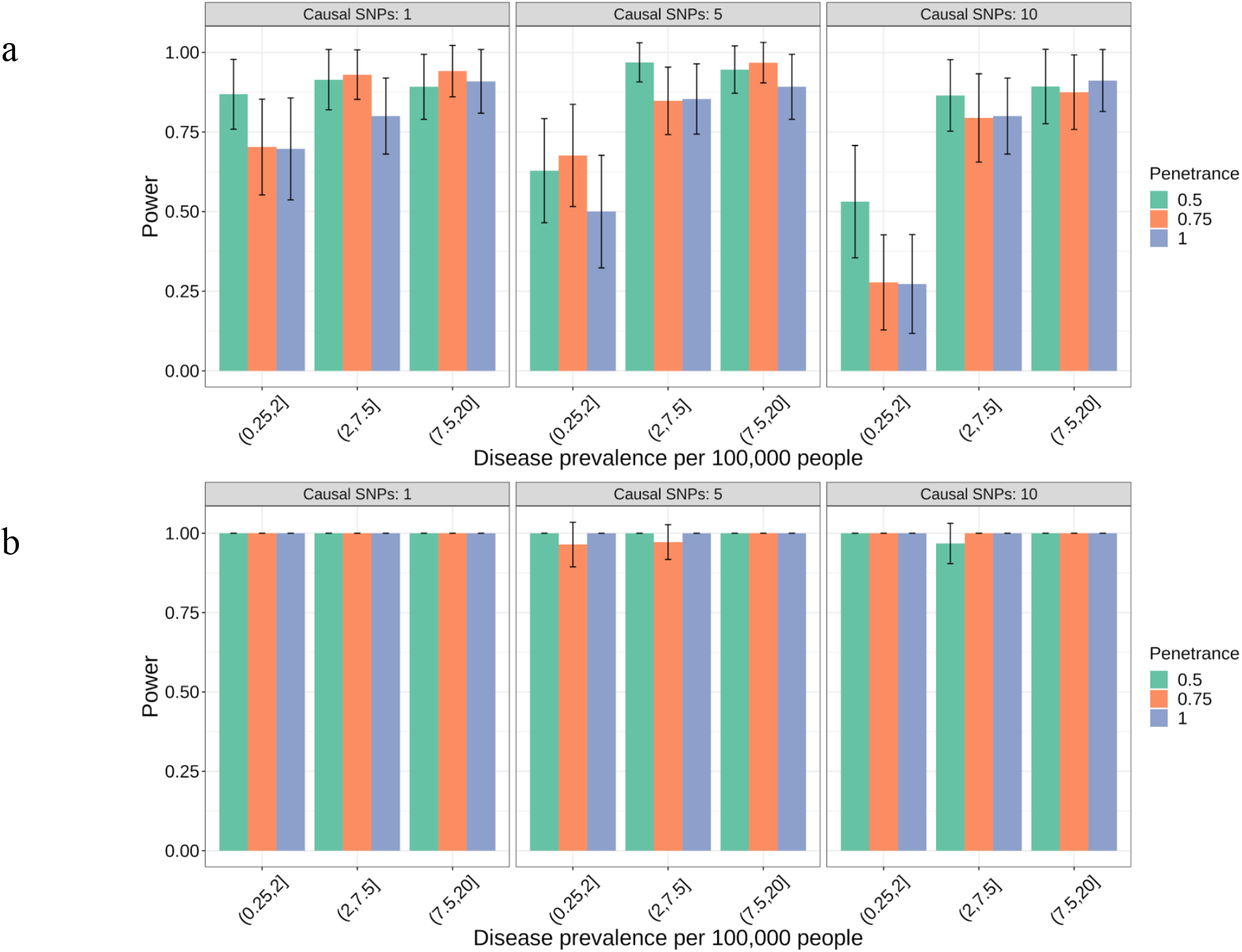
Power of GWAS to find associations at genome-wide significance (p < 5e-8) near causal genes for simulated rare diseases under (a) dominant inheritance model (b) recessive inheritance model.

#### Replication of rare disease associations

To verify if we could replicate known associations in rare diseases with the GWAS approach, we performed GWAS in individuals of European ancestry for 33 rare diseases. For each rare disease GWAS, cases were defined as having self-reported rare disease diagnosis in at least one online survey, and controls were defined as not having a self-reported rare disease diagnosis in any survey (Methods and Supplementary Notes Section 2). Table 1 shows the association statistics for GWAS hits from 18 rare diseases where our GWAS replicates associations near known genes at the genome-wide significance threshold, with no genome-wide significant associations found for the remaining 12 diseases. Supplementary Table S1 shows the case-control counts, number of GWAS hits and stratification correction for each GWAS, and Supplementary Figures 1-22 show the locuszoom plots for these associations. The lead variants for these associations span a wide range of minor allele frequencies (minimum MAF=0.0001, maximum MAF=0.4873) and effect sizes for the risk allele (minimum OR=1.2392, maximum OR=273.149). In addition, the associations were also found for a wide range of case counts (minimum number of cases=32, maximum number of cases=3,130), suggesting that GWAS in rare diseases can be performed with small case counts due to rare disease genetic architecture involving variants with large genetic effects. Of the diseases for which we replicated known associations, some appeared monogenic (Huntington’s disease, phenylketonuria, hemochromatosis, factor V deficiency), while others appeared polygenic (cleft lip, Waldenstrom macroglobulinemia, Hirschsprung’s disease), demonstrating the range of genetic architectures in which associations can be detected using GWAS.

**Table 1:** Discovery of known rare disease associations at genome-wide significance through GWAS on self-reported rare disease data.

| Phenotype | Cases | Lead SNP | P-value | RAF | OR | Nearby known gene/region |
| --- | --- | --- | --- | --- | --- | --- |
| Cystic fibrosis | 2482 | rs113993960 | 4.50e-305 | 0.01364 | 10.526<br>[9.615,11.628] | CFTR |
| Familial Mediterranean fever | 117 | rs200431216 | 1.50e-12 | 0.01060 | 7.460<br>[4.730,11.767] | MEFV |
| Class I glucose-6-phosphate dehydrogenase deficiency | 128 | rs180841877 | 1.80e-27 | 0.01226 | 8.714<br>[6.525,11.637] | G6PD |
| Beta Thalassemia | 771 | rs76053862 | 2.60e-105 | 0.00958 | 15.385<br>[12.658,18.868] | HBB |
| Lynch syndrome | 356 | rs565120523 | 3.50e-17 | 0.00087 | 31.112<br>[17.766,54.483] | MSH2, MSH6, EPCAM |
| Lynch syndrome | 356 | rs185663024 | 8.70e-12 | 0.00101 | 26.175<br>[13.641,50.226] | MSH2, MSH6 |
| Lynch syndrome | 356 | rs186191471 | 7.40e-09 | 0.00706 | 5.988<br>[3.690,9.709] | PMS2 |
| Primary myelofibrosis | 355 | rs10123976 | 2.90e-16 | 0.26399 | 1.933<br>[1.657,2.254] | JAK2 |
| Congenital factor V deficiency | 63 | rs6025 | 4.50e-68 | 0.03655 | 45.688<br>[30.941,67.465] | F5 |
| Hirschsprung disease | 79 | rs2435359 | 5.00e-26 | 0.25858 | 5.618<br>[4.049,7.813] | RET |
| Homocystinuria due to methylene tetrahydrofolate reductase deficiency | 70 | rs1801133 | 7.20e-12 | 0.34481 | 3.268<br>[2.309,4.630] | MTHFR |
| Huntington's disease | 41 | rs115335747 | 2.20e-08 | 0.04340 | 6.024<br>[3.571,10.204] | HTT |
| Phenylketonuria | 32 | rs4420324 | 2.10e-09 | 0.38742 | 5.127<br>[2.860,9.190] | PAH |
| Retinitis pigmentosa | 3130 | 3:128782923 | 2.10e-34 | 0.00010 | 273.149<br>[119.920,622.062] | RHO |
| Retinitis pigmentosa | 3130 | rs111033333 | 2.90e-16 | 0.00107 | 5.714<br>[4.065,8.000] | USH2A |
| Epidermolysis bullosa simplex | 35 | rs192032023 | 2.00e-09 | 0.00887 | 22.177<br>[10.640,46.222] | KRT5 |
| Cleft lip | 1581 | rs17461953 | 1.00e-11 | 0.19941 | 1.340<br>[1.234,1.455] | ARHGAP29 |
| Cleft lip/palate | 216 | rs113684606 | 1.30e-08 | 0.90217 | 5.065<br>[2.592,9.898] | MSX1 |
| Cleft lip | 1581 | rs2789341 | 1.90e-10 | 0.33194 | 1.277<br>[1.186,1.377] | PAX7 |
| Cleft lip | 1581 | rs563414765 | 2.50e-09 | 0.00129 | 5.464<br>[3.484,8.547] | 9q21 |
| Cleft lip | 1581 | rs34753522 | 9.80e-09 | 0.62539 | 1.245<br>[1.154,1.343] | MAFB |
| Cleft lip | 1581 | rs11078776 | 1.30e-08 | 0.55334 | 1.239<br>[1.151,1.335] | NTN1 |
| Cleft lip | 1581 | rs7069235 | 2.40e-08 | 0.16067 | 1.299<br>[1.188,1.420] | VAX1 |
| Cleft lip | 1581 | rs17242358 | 9.80e-37 | 0.19308 | 1.686<br>[1.560,1.821] | 8q24 |
| Corneal dystrophy | 314 | rs11659764 | 5.00e-30 | 0.05155 | 4.132<br>[3.344,5.102] | TCF4 |
| Craniosynostosis | 152 | rs1124471 | 9.10e-10 | 0.32104 | 2.062<br>[1.639,2.591] | BMP2 |
| Craniosynostosis | 152 | rs1894872 | 2.20e-08 | 0.48733 | 1.934<br>[1.527,2.445] | BBS9 |
| Waldenstrom macroglobulinemia | 90 | rs117972357 | 2.10e-14 | 0.00309 | 33.333<br>[17.544,62.500] | TCL1 |
| Waldenstrom macroglobulinemia | 90 | rs116446171 | 2.80e-11 | 0.01900 | 7.064<br>[4.444,11.228] | IRF4 |

#### *HTT* gene association in Huntington’s disease

Huntington’s disease is an autosomal dominant neurological disorder which manifests in adulthood with a combination of motor, cognitive and behavioural features^11^. The disease is caused by an expanded CAG trinucleotide repeat (of variable length) in *HTT*, the gene that encodes the protein huntingtin. We find a genome-wide significant association for Huntington’s disease at rs115335747 in the *HTT* gene (p=2.2e-8, MAF=0.0434, OR=6.024, Supplementary Figure 13) in our GWAS with 41 cases and 135,091 controls. The lead SNP rs115335747 is in linkage disequilibrium with rs149109767 (r-squared=0.56), a 3-bp deletion at codon 2642 in the *HTT* gene. The lead SNP and the linked coding indel have been reported in the literature as tagging variants for the major European risk haplotype (*hap*.*01*) for Huntington’s disease, occurring in nearly 50% of European Huntington’s disease cases^12^. The GWAS identifies rs115335747 as the lead SNP in the *HTT* gene with the risk allele matching that on the *hap*.*01* haplotype. While neither the lead SNP nor the linked coding variant are causal, their ability to tag a haplotype common in cases allows us to detect this association with a small number of cases.

#### Comparison of effect size estimates to published GWAS

We compared effect size estimates from GWAS on self-reported data to those from published GWAS (using either unrelated samples or case-parent trio designs) included in the GWAS catalog by matching hits based on linkage disequilibrium between lead variants (Methods). We found good agreement between effect size estimates for hits from GWAS on self-reported data to those included in the GWAS catalog, with overlapping confidence intervals for effect sizes of 4 out of 6 matched associations (Figure 2, Supplementary Table S5). The largest discrepancy between an effect-size estimate from self-reported data and GWAS catalog results is for Hirschsprung disease, where the reported effect in one of the studies in the GWAS catalog is from a recessive genetic model^13^. For Waldenstrom macroglobulinemia, the effect sizes reported in the GWAS catalog are from a cohort that is enriched for cases with family history of hematological malignancy^14^, which can inflate effect size estimates. However, when we compare our effect size estimates (risk allele = G, effect = 1.955, 95% CI= [1.492,2.418]) to the effect size from the replication cohort in the same study that includes only unrelated samples (risk allele = G, effect = 2.042, 95% CI = [1.495, 2.590]), we find the estimates nearly identical.

**Figure 2.**
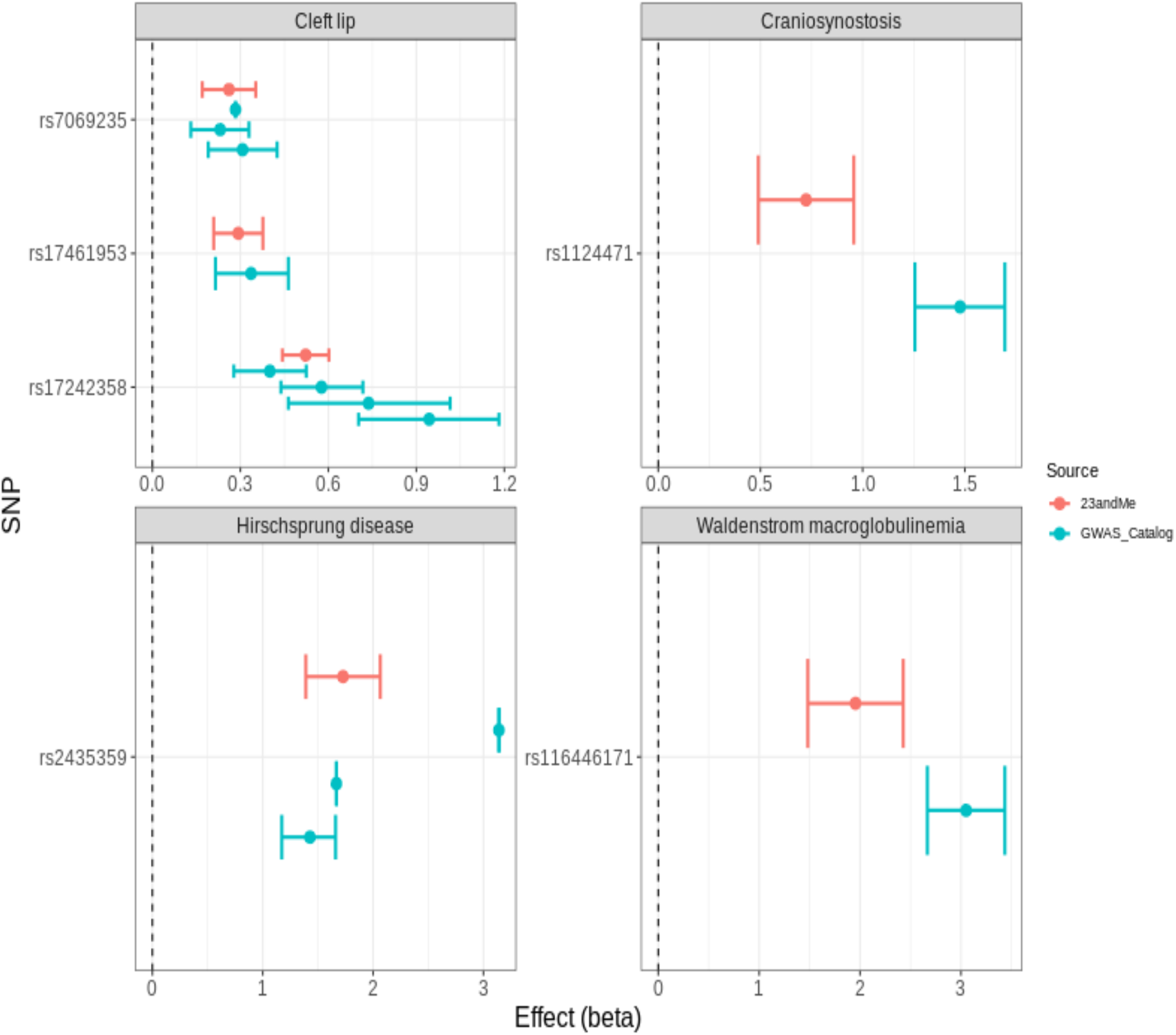
Comparison of effect size estimates between GWAS hits from self-reported data and matched GWAS catalog hits for the same phenotypes. Each panel shows a different rare disease, and rows within each panel indicate hits from the corresponding self-reported phenotype GWAS.

### Identification of novel associations

Having validated our approach, we next attempted to identify novel associations for rare diseases using GWAS on self-reported rare disease data. We found novel associations for four rare diseases - orofacial clefts, vestibular schwannoma, Duane retraction syndrome and spontaneous pneumothorax. We replicate these in the UK Biobank cohort, or in an independent subset of the 23andMe cohort when phenotypes were not available in the UK Biobank.

#### Orofacial cleft associations

Cleft lip with or without cleft palate is a rare disease that occurs at birth, with wide variability across geographic origin, racial and ethnic groups, as well as environmental exposures and socioeconomic status^15^. GWAS on cleft lip in Europeans and East Asians have identified a number of genetic associations^16^. Of these, we find associations at the genome-wide significance threshold for 8q24, 9q21, *PAX7, ABCA4, VAX1* and *NTN1* in our GWAS of cleft lip with 1,581 cases and 169,956 controls (Table 1).

We find a potentially novel association in the 3p22.1 region, at rs72419458 near the *POMGNT2* gene, in the GWAS for cleft lip (p=2.4e-8, MAF=0.072, OR=0.621, Supplementary Figure 23). Recently, this region has been reported to be associated with cleft palate only (i.e., cleft palate without cleft lip)^17^. Our GWAS results suggest that *POMGNT2* may also be associated with cleft lip.

#### Vestibular schwannoma association

Vestibular schwannomas are benign tumors of the Schwann cell sheath arising from the vestibular branch of the eighth cranial nerve^18^. Over ninety percent of vestibular schwannomas are unilateral^19^; bilateral vestibular schwannomas are primarily observed in patients with Neurofibromatosis Type 2, an autosomal dominant mendelian disorder, which is caused by inheritance of a germline coding mutation in the *NF2* gene on chromosome 22, encoding the tumor suppressor merlin/schwannomin. Mutations and copy number changes in *NF2* are the most frequent somatic changes observed in both unilateral and bilateral vestibular schwannomas^20,21^.

In our GWAS of vestibular schwannoma with 1,216 cases and 168,029 controls, we find one genome-wide significant hit at rs7341786 (p=1.4e-15, MAF=0.47437, OR=1.395, Figure 3), upstream of the *CDKN2A-CDKN2B* genes on chromosome 9, which control cell cycle checkpoints and DNA damage response pathways such as MDM2/p53. The *CDKN2A* and *CDKN2B* genes are tumor suppressor genes that have been previously associated with a variety of cancers, including germline susceptibility for familial atypical multiple mole melanoma (FAMM) syndrome^22^, meningioma^23^, colorectal cancer^24^, glioma^25^, acute lymphoblastic leukemia^26^, as well as somatic mutations in a number of additional cancer types^27^. A germline mutation in *CDKN2A*/*CDKN2B* has been associated with multiple nerve sheath tumors in a family with FAMM^28^, and loss of expression of these gene product(s) has been observed in schwannomas. However, the association between variant rs7341786 and vestibular schwannoma has not been reported previously, to our knowledge, and adds more evidence to the pleiotropic role of the *CDKN2A-CDKN2B* locus in cancer risk.

**Figure 3:**
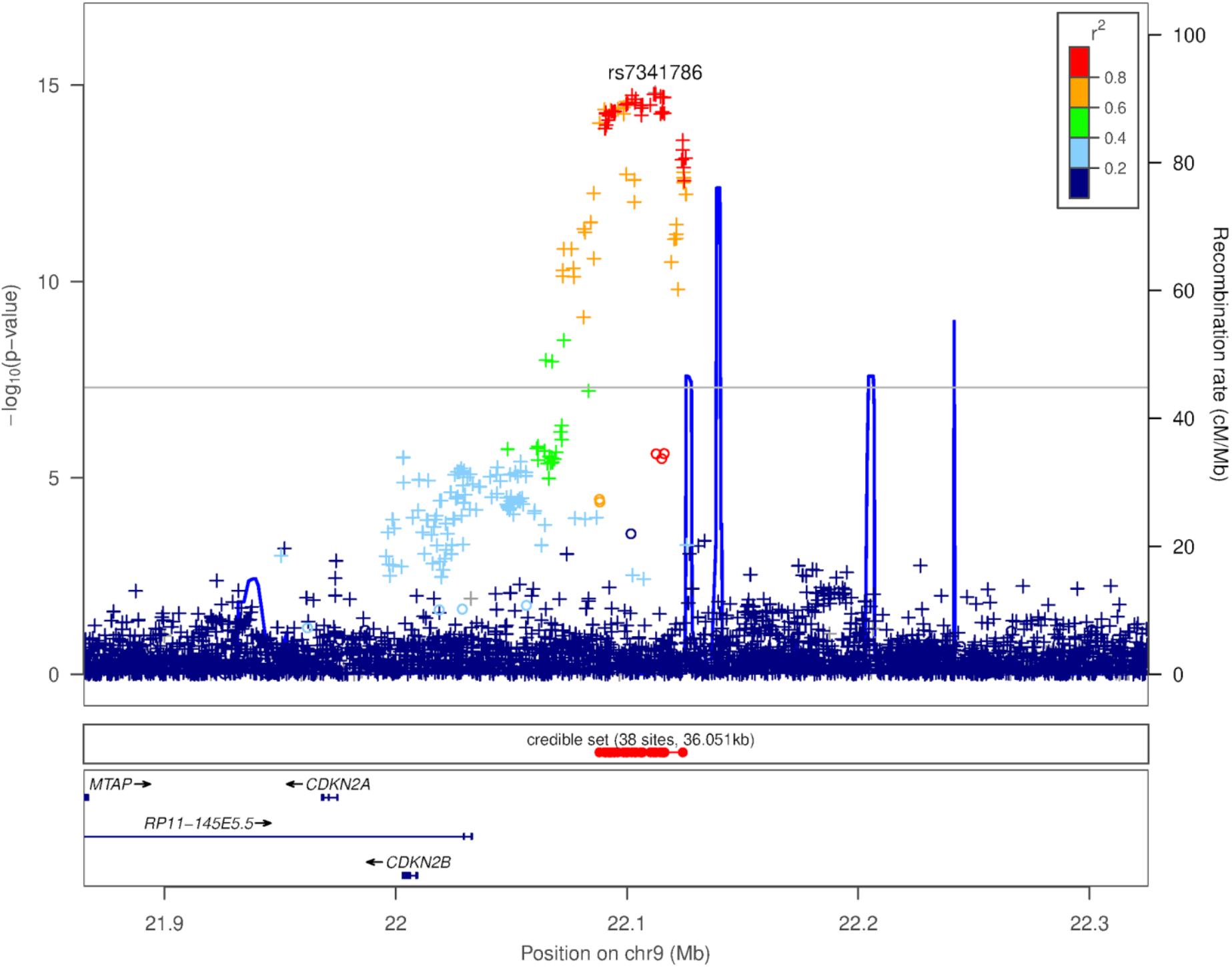
Locuszoom plot for vestibular schwannoma association

#### Duane retraction syndrome associations

Duane retraction syndrome is a congenital eye movement disorder, commonly characterized by problems with abduction (outward movement) of the eyes, and to a smaller extent, adduction (inward movement) of the eyes. It is believed to be caused by impaired development of motor neurons in the abducens nucleus and aberrant innervation of the lateral rectus muscle^29^. Mutations in 3 genes - *MAFB* (20q12)^30^, *SALL4* (20q13.2)^31,32^, and *CHN1* (2q31.1)^33^, have been associated with Duane retraction syndrome.

Our GWAS for Duane retraction syndrome includes 205 cases and 168,154 controls, and finds two genome-wide significant associations (1) rs9977152 (p=2.3e-14, MAF=0.27908, OR=2.378, Figure 4(a)) near *OLIG1* on chromosome 21 and (2) rs7834393 (MAF=0.00374,p=3.1e-08, OR=47.331, Supplementary Figure 24) near *UBE2W* on chromosome 8. In addition, we also find a conditional genome-wide significant hit, rs2105477 (p=2.1e-10 conditioned on rs9977152, MAF=0.44118, OR=1.908, Figure 4(b)), near *OLIG2*. This SNP is also marginally genome-wide significant (marginal p=4.07e-11). Of these, we suspect that the chromosome 8 association is likely to be a technical artifact based on lack of linkage disequilibrium to any other nearby SNPs (Supplementary Figure 26).

**Figure 4:**
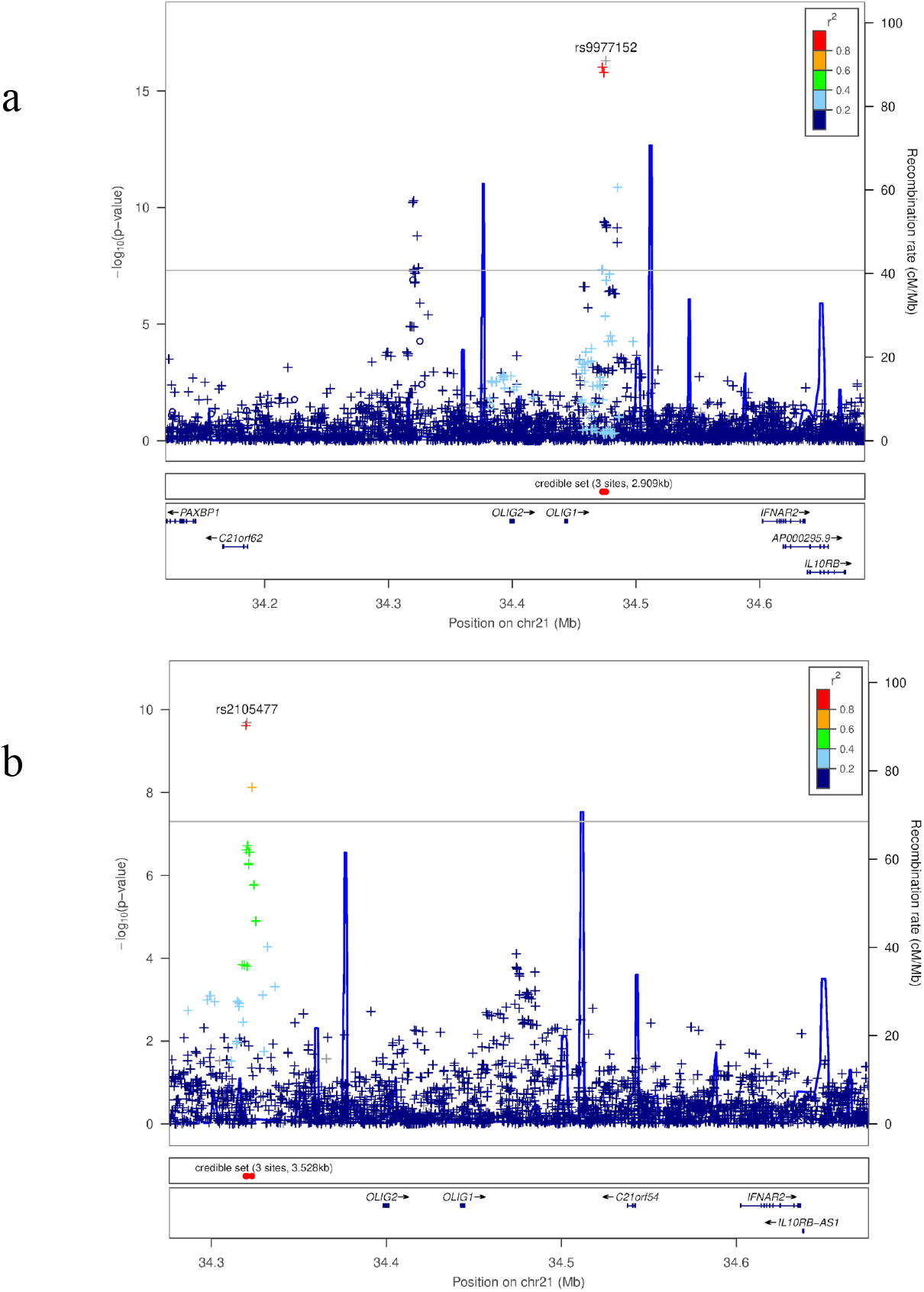
Locuszoom plot for duane retraction syndrome chromosome 21 associations (a) primary signal (b) conditional signal

The signal on chromosome 21 is not located near the known genetic causes of Duane retraction syndrome, suggesting a novel genetic association. The lead SNP, rs9977152, is an eQTL for the *OLIG1* gene in lung tissue in GTEx V8 (p=0.000080), with the T allele associated with decreased expression of *OLIG1* and increased risk for disease. Another independent signal at rs2105477, 155,475 bp away from the primary signal, was found through conditional analysis near the *OLIG2* gene. *OLIG1* and *OLIG2* are oligodendrocyte lineage transcription factors that are essential for oligodendrocyte development^34,35^. *OLIG2* is also essential for somatic motor neuron development in the spinal cord and hindbrain^36,37^. In zebrafish, *olig2* is necessary for the production of abducens motor neurons^38^. *OLIG2* expression in abducens motor neuron precursors is driven by *HOXA3* and *HOXB3*^39^, both of which are regulated by *MAFB*^40^, a known causal gene for Duane retraction syndrome^41^. *OLIG1* / *OLIG2* may thus contribute to risk of Duane retraction syndrome by affecting the development of motor neurons that control eye movements.

#### Spontaneous pneumothorax associations

Spontaneous pneumothorax is an abnormal accumulation of air in the space between the lungs and the chest cavity that can result in the partial or complete collapse of a lung.

Spontaneous pneumothorax is classified as primary when it occurs without underlying lung disease, and as secondary when underlying respiratory diseases such as emphysema or asthma, acute or chronic infections, lung cancer are present^42^. Mutations in the *FLCN* gene (17p11.2) have previously been associated with primary spontaneous pneumothorax^43–45^.

We conducted a GWAS for spontaneous pneumothorax with 1,143 cases and 168,042 controls, and found three potentially novel associations. First, we find a genome-wide significant association at rs4437193 in the *KY* gene on chromosome 3 (p=3.7e-10, MAF=0.38699, OR=0.7535, Figure 5). This association is in linkage disequilibrium with two splice variants in the adjacent *CEP63* gene (rs35934324 with r-squared = 0.75, and rs11927068 with r-squared =0.74). The lead SNP rs4437193 is also an eQTL for the *ANAPC13* gene in 43 tissues including lung tissue (p=2.3e-55) and for *CEP63* in 35 tissues including lung tissue (p=1.7e-29) in GTEx V8. Therefore the causal gene and mechanism of action for this association is unclear.

**Figure 5:**
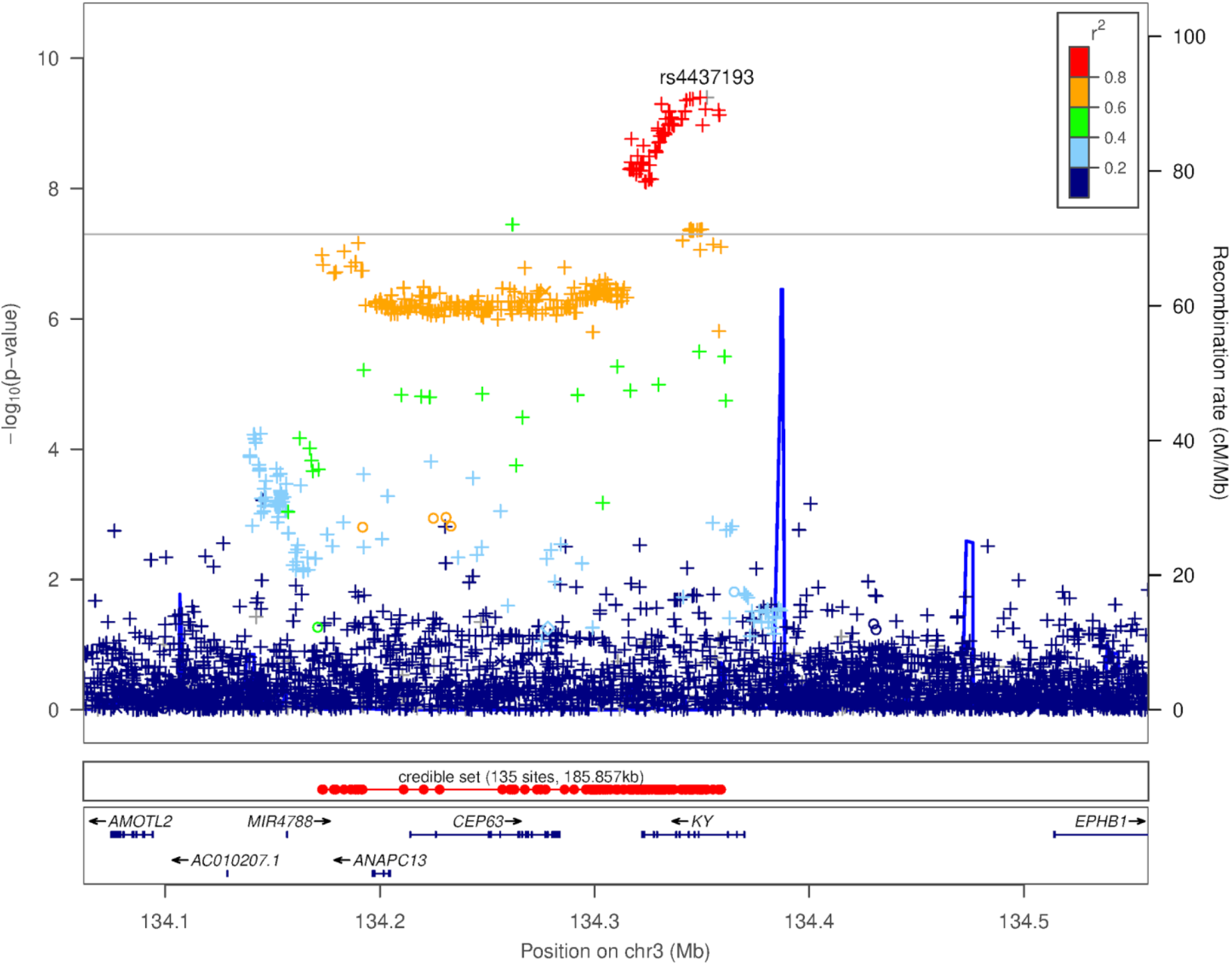
Locuszoom plot for spontaneous pneumothorax chromosome 3 association

We also find a second genome-wide significant association at rs9547906 near the periostin gene *POSTN* on chromosome 13 (p=7.8e-10, MAF=0.3357, OR=1.310, Figure 6). The lead SNP rs9547906 is an eQTL for the *POSTN* gene in 3 tissues (Artery - Aorta, p = 4.4e-10; Colon - Sigmoid, p = 3.9e-7; Pancreas, p = 0.0000064) and for the *TRPC4* gene in 1 tissue (Adipose - Subcutaneous, p = 0.00012). In addition, the lead SNP is also in high LD (r-squared = 0.99) with esv3631801, a 13,638-bp deletion. Periostin, the protein produced by the *POSTN* gene, is known to be involved in airway development^46^ and plays an important role in the cell cycle of lung fibroblasts^47^. Periostin also plays an important role in other respiratory diseases including asthma^48^ and idiopathic pulmonary fibrosis^49^. Our GWAS suggests that periostin may also contribute to susceptibility for spontaneous pneumothorax.

**Figure 6:**
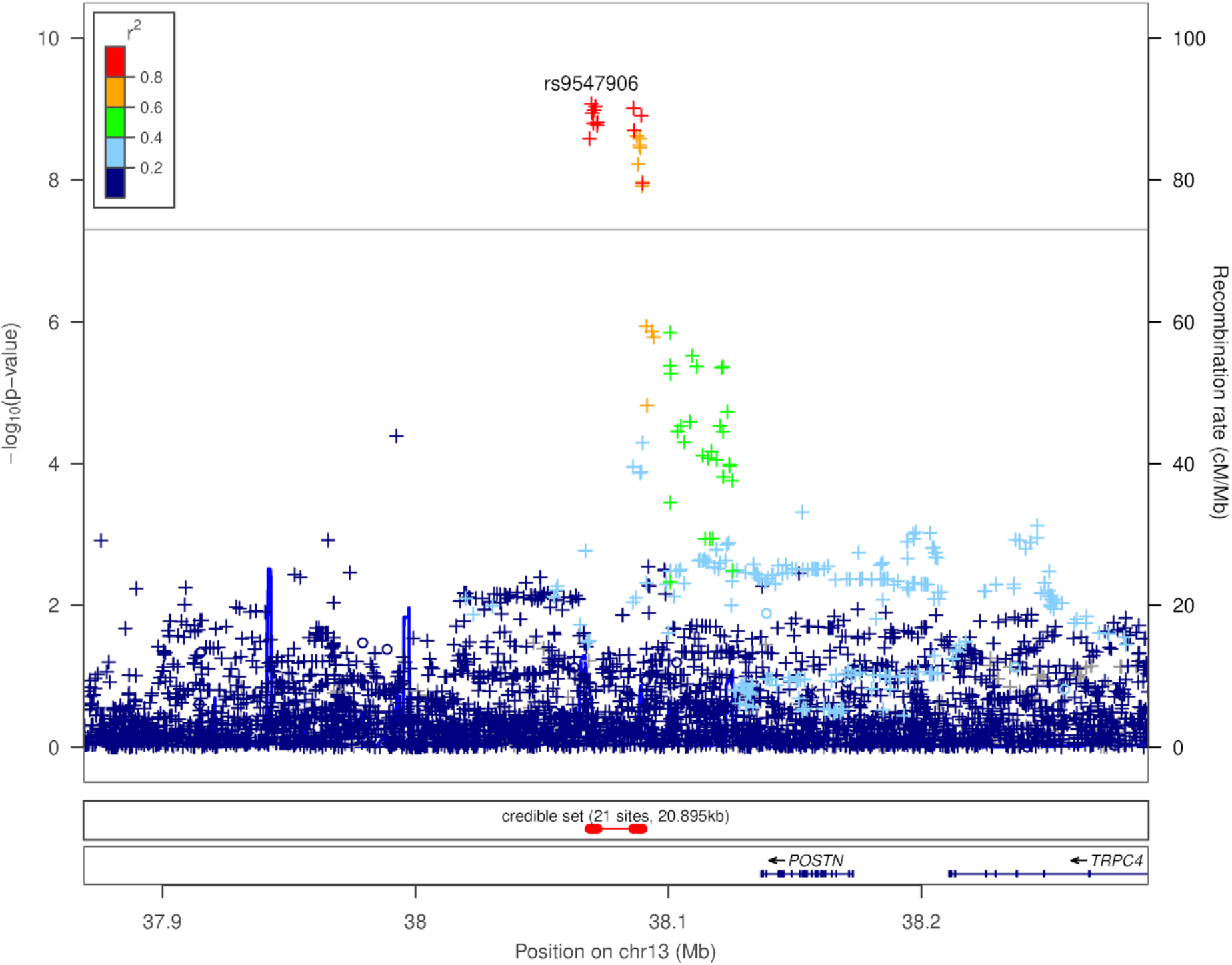
Locuszoom plot for spontaneous pneumothorax chromosome 13 association

A third genome-wide significant association is found at rs2161648 on chromosome 16 (p=4.8e-08, MAF=0.41318, OR=1.268, Figure 7) near the *BCAR1* and *CFDP1* genes. The lead SNP is an eQTL for multiple nearby genes, including *BCAR1* (7 tissues), *CFDP1* (12 tissues) and *TMEM170A* (8 tissues). Variants linked to our lead SNP have previously been reported to have associations with lung-function phenotypes (FEV1, FEV1/FVC ratio, peak expiratory flow^50^) as well as chronic obstructive pulmonary disease^51,52^. We find that the lead SNP is in high LD (r-squared = 0.93) with esv3639091, an 843-bp intronic deletion in *CFDP1*, that has been reported to be an eQTL for *CFDP1, BCAR1, ADAT1*, and *RFWD3* in an joint SNP-SV analysis of gene expression in GTEx data^53^.

**Figure 7:**
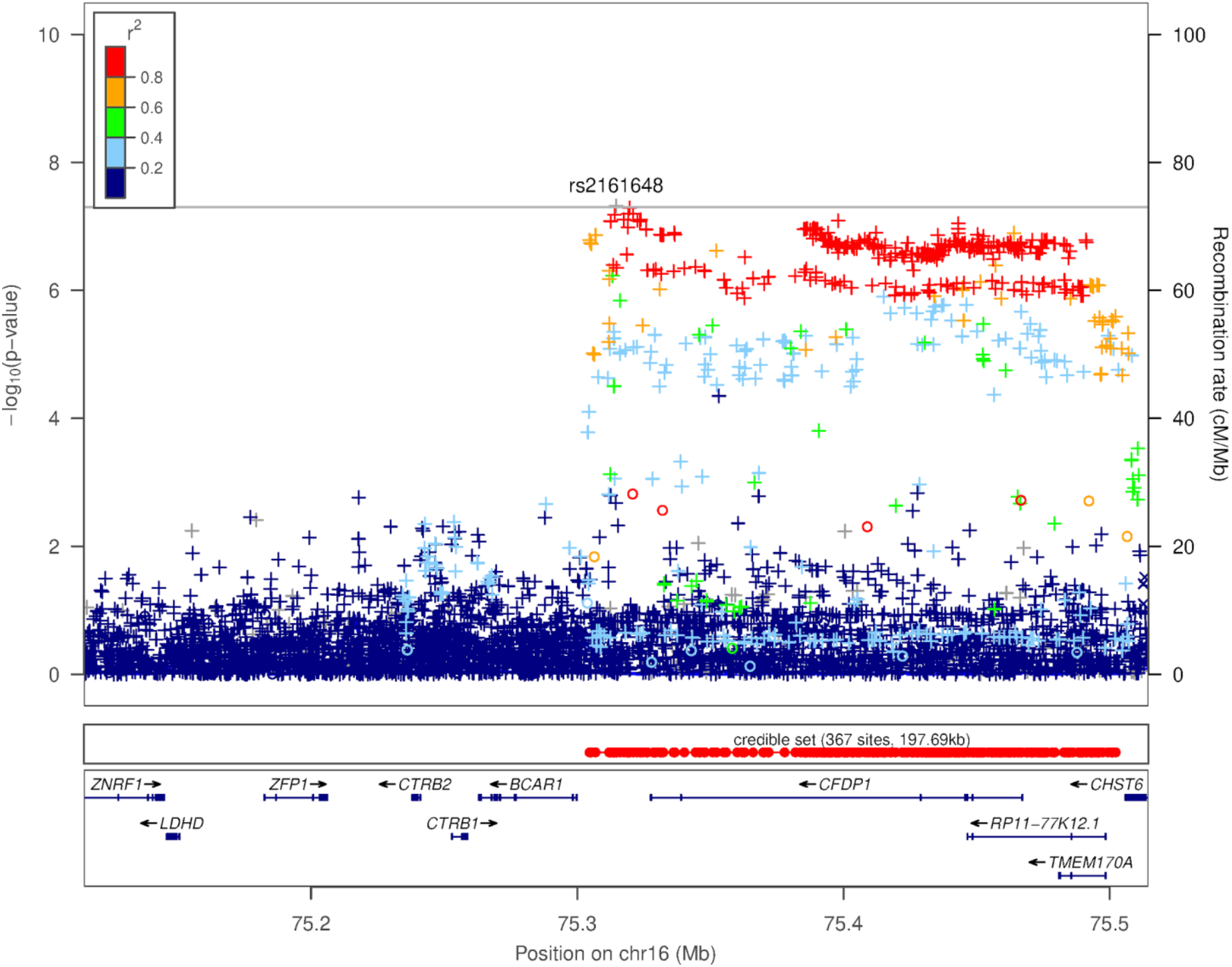
Locuszoom plot for spontaneous pneumothorax chromosome 16 association

#### Replication of novel associations

We attempted to replicate the novel associations we discovered using the UK Biobank cohort. Where phenotypes were not available in the UK Biobank, we attempted replication in a cohort of consented 23andMe research participants who were not included in the discovery GWAS. We find that 3 of the 6 associations replicate at p < 0.05, and 5 out of 6 show the same direction of effect in the replication and discovery cohorts (Table 2, Supplementary Table S6).

**Table 2:** Replication of novel associations. We performed replication in the UK Biobank dataset, and where phenotypes were not available in the UK Biobank we used a cohort of research-consented 23andMe customers who were not included in the discovery GWAS.

| Phenotype | Lead SNP | Replication cohort | Discovery p-value | Replication p-value | Replication cases | Effect direction match |
| --- | --- | --- | --- | --- | --- | --- |
| Vestibular schwannoma | rs7341786 | UKBB | 1.4e-15 | **1.56e-2** | 232 | True |
| Spontaneous pneumothorax | rs4437193 | UKBB | 3.7e-10 | 9.69e-1 | 707 | True |
| Spontaneous pneumothorax | rs9547906 | UKBB | 7.8e-10 | **5.29e-3** | 707 | True |
| Spontaneous pneumothorax | rs2161648 | UKBB | 4.8e-08 | 2.48e-1 | 707 | True |
| Duane retraction syndrome | rs9977152 | 23andMe | 3.5e-17 | **7.01e-06** | 44 | True |
| Cleft lip | rs72419458 | 23andMe | 2.4e-8 | 8.06e-01 | 310 | False |

### Trans-ethnic analyses

To study rare diseases in all populations, we attempted to replicate known rare disease associations in non-European populations. Since the sample size for rare diseases in non-Europeans is smaller than Europeans in the 23andMe cohort, we also performed trans-ethnic analyses including data from all populations using mixed-models.

#### Trans-ethnic replication of known rare disease associations

To better understand the genetic architecture of rare diseases across populations, we further attempted to replicate the significant known associations from European GWAS in each of the African-American, East Asian, Latino, and South Asian populations, when we had sufficient cases. Table 3 summarizes association results for class I glucose-6-phosphate dehydrogenase deficiency (G6PD deficiency) and beta thalassemia for populations where we have more than 30 cases.

**Table 3.** Trans-ethnic replication of European hits

| Phenotype | Ethnicity | Cases | SNP | P-value | MAF | OR |
| --- | --- | --- | --- | --- | --- | --- |
| Class I glucose-6-phosphate dehydrogenase deficiency | African American | 214 | rs150053350 | 2.1e-70 | 0.13 | 4.92 [4.09, 5.93] |
|  | East Asian | 74 | rs142551997 | 1.85e-03 | 0.04 | 15.06[0.76,298.5] |
|  | Latino | 94 | rs150053350 | 1.28e-42 | 0.012 | 9.37 [7.12, 12.33] |
| Beta thalassemia | African American | 30 | rs76053862 | 2.06e-01 | 0.004 | 6.97 [0.65, 75.26] |
|  | East Asian | 52 | rs34451549 | 2.43e-14 | 0.002 | 58.56 [24.05, 142.57] |
|  | Latino | 45 | rs76053862 | 1.06e-06 | 0.007 | 21.03 [8.50, 52.01] |
| Craniosynostosis | Latino | 42 | rs1124471 | 1.98e-4 | 0.37 | 2.33 [1.49,3.67] |

G6PD is a common disease that affects 11-13% of African-Americans^54^. In the African-American association analysis, although the lead SNP from the European GWAS (rs180841877) failed quality control, we identified a different SNP, rs150053350, 15,490-bp away, that reached genome-wide significance (p=2.1e-70, MAF=0.13, OR=4.92). This SNP is also the most significant SNP in the Latino analysis (p=1.28e-42, MAF=0.012, OR=9.37).

Although the number of cases of beta thalassemia in the non-European population is relatively small, the lead SNP from the European GWAS was nominally significant in Latino association analysis (p=1.6e-6, MAF=0.007, OR=21.03). In the East Asian association analysis, rs34451549, 121,171-bp away from the European lead SNP, reached genome-wide significance (p=2.43e-14, MAF=0.002, OR=58.56). We also replicated the craniosynostosis association for rs1124471 at nominal significance in the Latino analysis with 42 cases (p=1.98e-4, MAF=0.37, OR=2.33).

#### Trans-ethnic mixed-model analysis of rare diseases

For phenotypes with potential novel association findings (cleft lip, vestibular schwannoma, Duane retraction syndrome, spontaneous pneumothorax) in the European GWAS, we further performed trans-ethnic mega GWAS with individual level data across all ethnicities using mixed model analyses implemented in SAIGE^55^. Table 4 indicates association statistics for lead variants in loci that reached genome-wide significance level in the trans-ethnic GWAS. The number of participants across ethnicities for each phenotype can be found in the Supplementary Table S4.

**Table 4.**
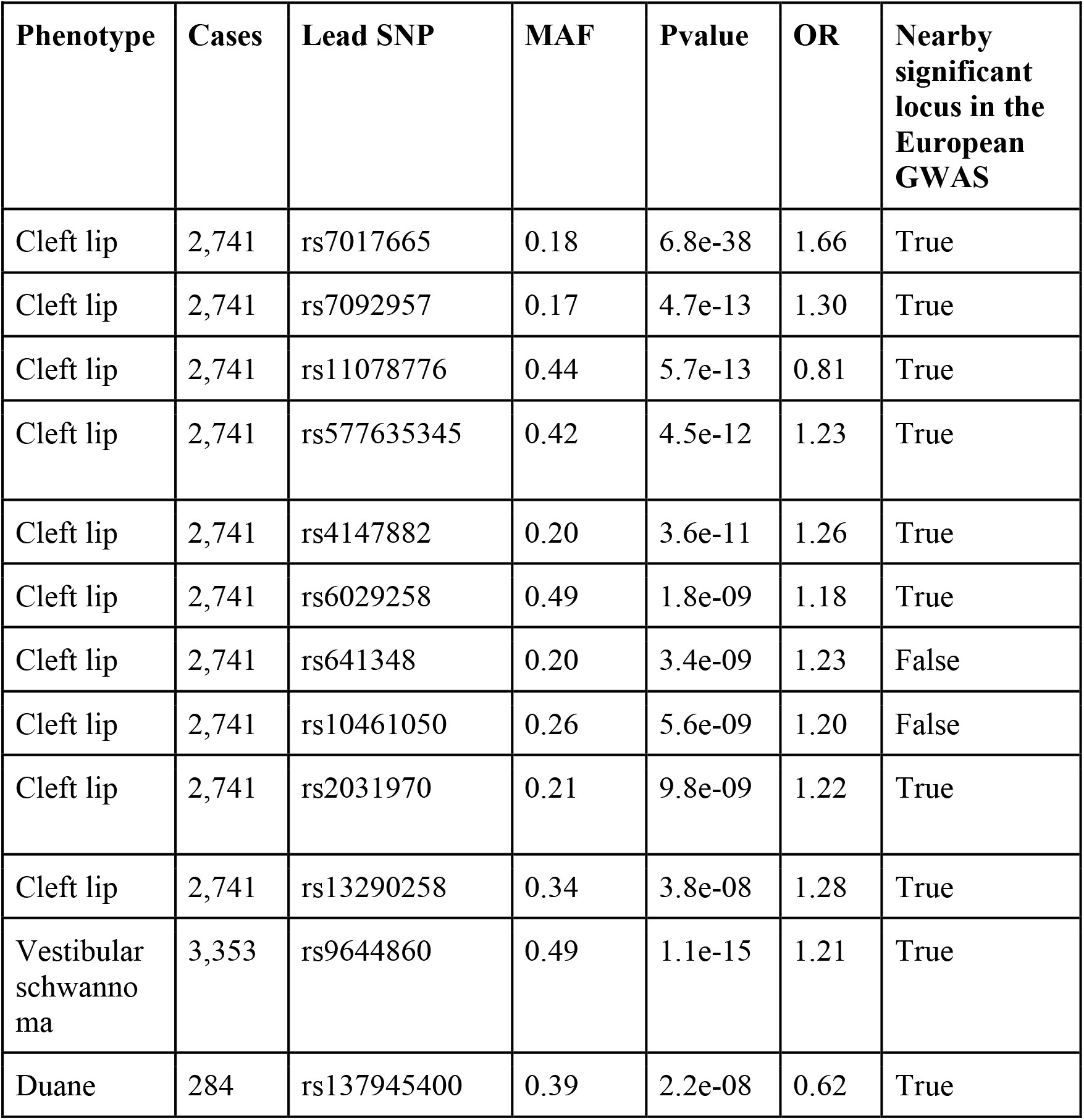
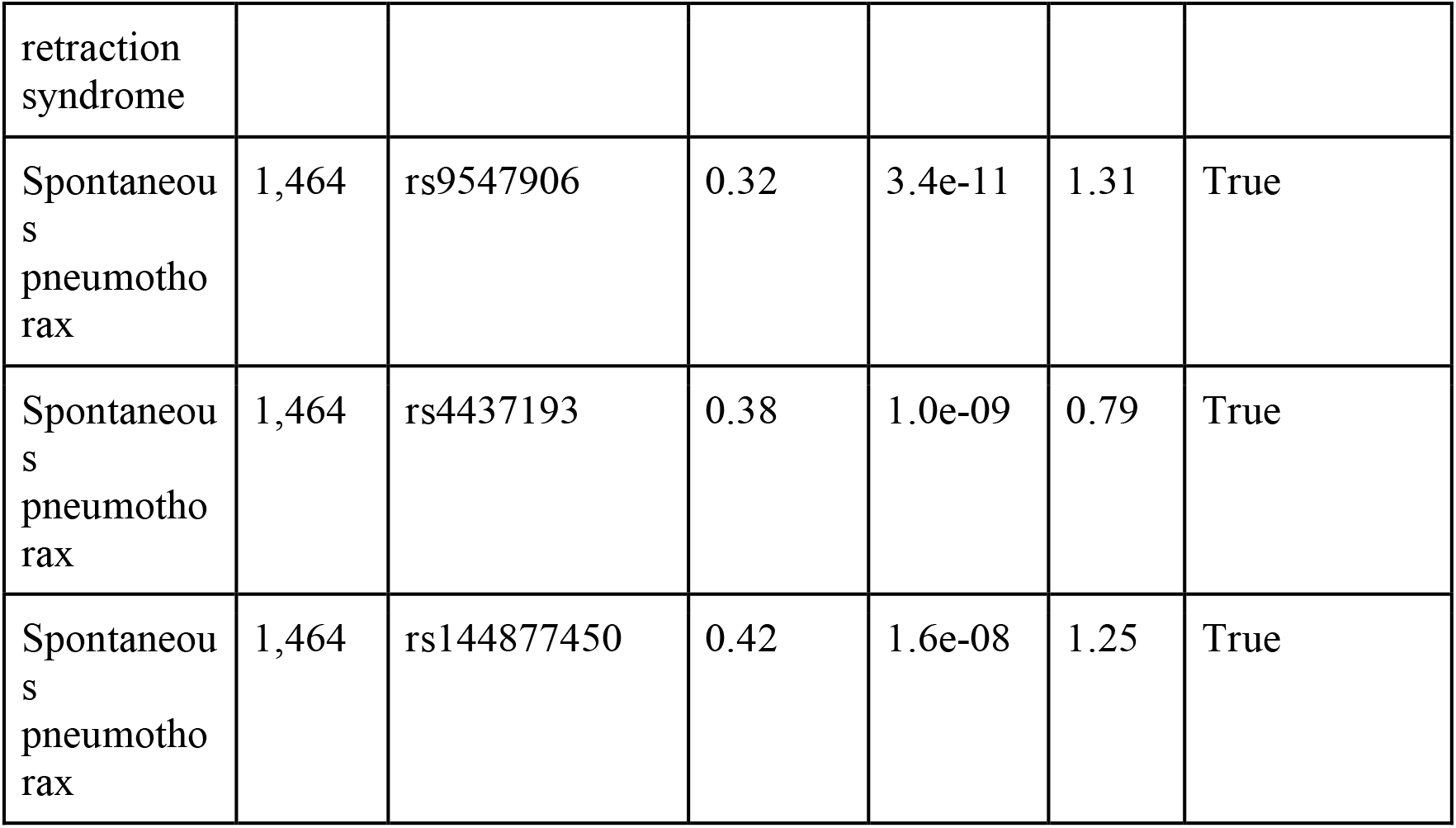
Significant loci in the trans-ethnic mega GWAS

A total of 2,741 cases and 241,298 controls were included in the trans-ethnics GWAS for cleft lip. In addition to loci that reach genome-wide significance level in the European GWAS, two additional loci reached genome-wide significance level in the trans-ethnic GWAS: one near the *IRF6* gene, one near the *DLGI* gene (Supplementary Figures 25 and 26). These two regions have been reported to be associated with cleft lip/palate in previous studies^56,57^. The potential novel association in the European GWAS (rs72419458) was not replicated in the trans-ethnic GWAS. The leading SNP p-value for other significant loci are more significant in the trans-ethnic GWAS compared to the European GWAS, largely due to the increased sample size.

Novel association findings in the European GWAS of vestibular schwannoma, Duane retraction syndrome and spontaneous pneumothorax all reached genome-wide significance in the trans-ethnic GWAS. P-values in the trans-ethnic GWAS are comparable or slightly more significant than the European GWAS.

## Discussion

A major challenge in the study of rare diseases is the difficulty of assembling sufficiently large case cohorts. Here, we demonstrate that self-reported rare disease diagnosis data can be used to study rare diseases at scale. To our knowledge, this is the largest study of rare diseases using genomic data, including 19,084 cases from 7 populations across 33 rare diseases. We ran GWAS for rare diseases using self-reported rare disease diagnosis. For some of these diseases, our study is the first GWAS, often replicating associations known through sequencing of cases or case families. Our GWASes of vestibular schwannoma, Duane retraction syndrome, and spontaneous pneumothorax are the first GWASes of these diseases that report novel genome-wide significant associations.

We validated our approach by replicating known rare disease associations through GWAS on self-reported rare disease data. We were able to re-discover 29 known associations at the genome-wide significance level. The replicated associations include associations for monogenic diseases as well as polygenic diseases. For additional validation, we compared effect sizes for genome-wide significant variants for rare diseases with published GWAS summary statistics. We found good agreement of effect sizes from GWAS on self-reported rare disease data with published effect sizes from clinical cohorts.

Given the wide variety of genetic architectures for monogenic diseases, with the number of causal variants in a gene ranging from one (e.g Huntington’s disease) to hundreds (e.g phenylketonuria), the ability of GWAS to find associations at rare causal genes for monogenic diseases in an outbred population is to some extent unexpected. In Huntington’s disease, we find a genome-wide significant association with 41 cases at rs115335747, a SNP that tags a risk haplotype (hap.01) that is common in European cases. Similarly, in phenylketonuria, we find a common association (MAF=0.38742) with 32 cases near the *PAH* gene that is casual for the disease. Many haplotypes of the *PAH* gene are known to be present at increased frequencies in phenylketonuria cases^58–60^. Since these haplotypes are based on restriction fragment length polymorphisms, we were unable to link our GWAS signal to any of these haplotypes. To examine the power of GWAS to find associations in or near causal genes in rare monogenic diseases, we simulated rare diseases in a cohort of 4,957,230 individuals of European ancestry. We found that GWAS is well-powered to find associations in both dominant and recessive monogenic diseases for a variety of architectures. In both settings, we found that GWAS lead SNPs have higher frequency than the causal variants, suggesting that GWAS cannot directly identify causal variants but can identify more common tag SNPs for the causal variants. A consequence of this is that in dominant rare diseases, GWAS has more power for less penetrant diseases than highly penetrant diseases, since the causal variants are more frequent in the low-penetrance setting.

We found 6 novel associations for 4 rare diseases - cleft lip, vestibular schwannoma, Duane retraction syndrome, and spontaneous pneumothorax. 5 of 6 showed the same direction of effect in a separate cohort of either UK Biobank or 23andMe research participants, and 3 of 6 replicated at p < 0.05. These associations are relatively common in frequency and are not located near known genetic causes of the respective diseases. This suggests that they are either modifiers for the known disease-associated genes or contribute independent risk. Recent studies have shown that penetrance of rare monogenic variants can be modified by a polygenic background^61,62^. Alternatively, epistatic interactions between rare variants and common variants can lead to increased susceptibility for rare disease^63–66^.

Our GWAS for Duane retraction syndrome is the first GWAS for the disease and we discovered two independent associations near the *OLIG1* and *OLIG2* genes, with eQTL evidence linking the first association to *OLIG1*. The role of *OLIG1* and *OLIG2* in oligodendrocyte and motor neuron formation is well-established^34–37^, and knockdown of *olig2* causes a lack of abducens motor neurons in zebrafish^38^. In addition, *OLIG2* may act downstream of *MAFB*, a known causal gene for Duane retraction syndrome^41^. This suggests that *OLIG1*/*OLIG2* contribute to susceptibility for Duane retraction syndrome, though it is unclear whether this risk is independent of known causal genes. It has been hypothesized that *OLIG1* and *OLIG2* may be involved in neurological diseases. Earlier genetic studies in schizophrenia have found associations for SNPs in *OLIG2*^67,68^, however, these results have not replicated in a larger recent schizophrenia GWAS^69^. There is evidence from human organoid and mouse models that *OLIG2* contributes to abnormal neurodevelopmental phenotypes in Down Syndrome^70,71^. Our GWAS provides human genetics evidence supporting the role of the *OLIG1* and *OLIG2* genes in neurological diseases.

We found 3 novel associations in our GWAS of spontaneous pneumothorax. Of the associations we found, 2 regions have been previously reported to be associated with lung function phenotypes. We also found that two lead variants were in high LD with structural variants (deletions in both cases), and one of the linked structural variants is an eQTL for multiple tissues in GTEx data. Structural variants have been causally linked to many common and rare diseases and are believed to act by affecting gene expression^72^.

Due to the composition of the 23andMe research participant base, we had the largest sample size for analysis in populations of European ancestry. Where sufficient sample size (at least 30 cases) were available in African-American, East Asian, Latino and South Asian populations, we attempted to replicate known and novel associations we had observed in European populations. We find that similar to GWAS replication for common diseases, trans-ethnic analyses show associations in the same genomic region but occasionally with different lead SNPs, reflecting differences in allele frequencies^73^ and linkage disequilibrium^74^. Similar to the disparities observed in GWAS studies of common diseases^75^, the relatively smaller numbers of research participants from non-European populations result in few rare diseases that could be analyzed across all ethnicities with sufficient power using a standard GWAS approach. To address this problem, we ran mixed-model analyses that included related and unrelated participants from all ethnicities. We found that trans-ethnic analyses had higher power than European GWAS. For rare diseases, where small samples sizes are often limit analyses, trans-ethnic mixed-model analyses can be an effective way of aggregating data.

Our analysis included as cases only those samples who self-reported a diagnosis of a rare disease. Therefore, errors in the original diagnosis or the self-reporting could affect our GWAS results. We expect that our use of the autocomplete feature in the rare disease survey, instead of thousands of “yes/no” questions for all known rare diseases, would allow more accurate self-reporting of diagnoses (Methods and Supplementary Notes). Our GWAS only included genotyped or well-imputed SNPs, indels and structural variants. Therefore, our analysis may not be well-powered for identifying associations if there is a single underlying rare causal variant that is poorly imputed or not assayed. Alternatively, if many different rare causal variants in a single gene lie on different haplotypes, none of the individual haplotypes may be imputed well enough to be associated in the GWAS but could be discovered through analysis of families or sequencing of unrelated cases. In simulations,we find that as disease prevalence decreases and the number of causal variants increases, the power of GWAS to find associations for dominant diseases decreases (Figure 1). An example of the limits of the GWAS for discovering associations in rare disease can be found in our analyses of vestibular schwannoma, Duane retraction syndrome, and spontaneous pneumothorax. For these diseases, our GWAS finds novel genetic associations, but fails to find associations at known causal genes, some of which have been found through sequencing of affected families^30,33^.

Despite these limitations, our results show that self-reported rare disease data is a viable method for discovering genetic associations for rare diseases. GWAS on rare disease data is a complementary approach to sequencing for the study of rare disease genetics since GWAS is well-powered for finding common variant associations to disease susceptibility. The results from our GWAS analyses show that rare diseases thought to be Mendelian may also have disease susceptibility attributable to common genetic variants. A promising future direction for studying rare diseases with self-reported data is the ability to collect information on family history for rare diseases. Genome-wide association studies by proxy, which replace cases by their first degree relatives, have been shown to improve power for diseases with low prevalence^76^. Haplotype association, rather than single-SNP association, could also be used to understand whether associations found through GWAS are capturing contributions of rare variants through common SNPs. With increasing sample size and diverse imputation reference panels, we may also be able to study rare diseases more widely in multiple populations and improve our understanding of the trans-ethnic genetic architecture of these diseases. These improvements can lead to increased genetic discoveries in rare diseases and potentially accelerate the path to finding therapies to cure them.

## Methods

### Subjects

Participants provided informed consent and participated in the research online, under a protocol approved by the external AAHRPP-accredited IRB, Ethical & Independent Review Services (E&I Review). Participants were included in the analysis on the basis of consent status as checked at the time data analyses were initiated.

Over 1.6 million genotyped participants were included in analyses based on selection for completing surveys about their health history and diagnosis of rare diseases.

### Self-reported phenotype data collection for rare diseases

We collected self-reported data on rare disease diagnosis history using an online survey. Disease names were ascertained from a list of 9000+ rare disease terms from Orphanet (accession: April 2017), and any diseases that could not be mapped to the list were recorded as ‘other’. Participants were asked to enter at least the first three letters of the rare condition(s) they have been diagnosed with, and a list of matching entries appears from which participants could select their response. The full list of diseases is available as Supplementary Table S1. For some rare diseases, health history surveys and questions also contained yes/no questions to ascertain disease diagnosis. For these diseases, a subject was defined as a case if they had indicated in either of the surveys that they had been diagnosed with the disease. More details on the questions and combination logic is available in Supplementary Notes Section 1. Case-control counts for each phenotype are available in Supplementary Table S2.

### Genotyping and SNP imputation

DNA extraction and genotyping were performed on saliva samples by CLIA-certified and CAP-accredited clinical laboratories of Laboratory Corporation of America. Samples were genotyped on one of five genotyping platforms. The v1 and v2 platforms were variants of the Illumina HumanHap550+ BeadChip, including about 25,000 custom SNPs selected by 23andMe, with a total of about 560,000 SNPs. The v3 platform was based on the Illumina OmniExpress+ BeadChip, with custom content to improve the overlap with our v2 array, with a total of about 950,000 SNPs. The v4 platform was a fully customized array, including a lower redundancy subset of v2 and v3 SNPs with additional coverage of lower-frequency coding variation, and about 570,000 SNPs. The v5 platform, in current use, is an Illumina Infinium Global Screening Array with approximately 640,000 SNPs) supplemented with approximately 50,000 SNPs of custom content. Samples that failed to reach 98.5% call rate were re-analyzed. Individuals whose analyses failed repeatedly were re-contacted by 23andMe customer service to provide additional samples.

### Imputation

Variants were imputed in two separate imputation reference panels. For the first panel, we combined the May 2015 release of the 1000 Genomes Phase 3 haplotypes^77^ with the UK10K imputation reference panel^78^ to create a single unified panel. To do this, multiallelic sites with N alternate alleles were split into N separate biallelic sites. We then removed any site whose minor allele appeared in only one sample. For each chromosome, we used Minimac3^79^ to impute the reference panels against each other, reporting the best-guess genotype at each site. This gave us calls for all samples over a single unified set of variants. We then joined these together to get, for each chromosome, a single file with phased calls at every site for 6,285 samples. Throughout, we treated structural variants and small indels in the same way as SNPs. We used the Human Reference Consortium (HRC) as the second imputation reference panel^80^. It consists of 32,488 samples for 39,235,157 SNPs.

In preparation for imputation we split each chromosome of the reference panel into chunks of no more than 300,000 variants, with overlaps of 10,000 variants on each side. We used a single batch of 10,000 individuals to estimate Minimac3 imputation model parameters for each chunk.

To generate phased participant data for the v1 to v4 platforms, we used an internally-developed tool, Finch, which implements the Beagle graph-based haplotype phasing algorithm^81^, modified to separate the haplotype graph construction and phasing steps. Finch extends the Beagle model to accommodate genotyping error and recombination, in order to handle cases where there are no consistent paths through the haplotype graph for the individual being phased. We constructed haplotype graphs for all participants from a representative sample of genotyped individuals, and then performed out-of-sample phasing of all genotyped individuals against the appropriate graph. For the X chromosome, we built separate haplotype graphs for the non-pseudoautosomal region and each pseudoautosomal region, and these regions were phased separately. For the 23andMe participants genotyped on the v5 array, we used a similar approach, but using a new phasing algorithm, Eagle2^82^.

We imputed phased participant data against both reference panels using Minimac3, treating males as homozygous pseudo-diploids for the non-pseudoautosomal region.

We then built a merged imputation dataset by combining the two sets of imputed data. We applied a simple merging rule: if a variant was imputed in the HRC panel, the HRC imputed results were included in the merged imputed dataset. For the remaining variants not present in HRC (all INDELs and structural variants, for example), the 1KG/UK10K imputed results were added to the merged dataset. In total, the merged imputed dataset contained 64,439,130 variants.

### Genome-wide association analysis

We selected case-control phenotypes with 30 or more cases for GWAS analysis. We performed GWAS on Europeans, Latinos, African-Americans, East Asians and South Asians separately. Individuals were assigned to ancestry groups based on a local-ancestry method^83^. Since most rare diseases had fewer than 10,000 cases but millions of controls, we downsampled controls to use only 5% of the total controls at random.

For each study, a maximal set of unrelated individuals was chosen using a segmental identity-by-descent estimation algorithm^84^. Individuals were defined as related if they shared more than 700 cM of identity-by-descent, including regions where the two individuals share either one or both genomic segments identity-by-descent. If a case and a control were identified as IBD with each other, we preferentially excluded the control from the study.

We computed association test results for the genotyped and the imputed SNPs by logistic regression assuming additive allelic effects. For tests using imputed data, we used the imputed dosages rather than best-guess genotypes. We included covariates for age, gender, the top five principal components to account for residual population structure, and indicators for genotype platforms to account for genotype batch effects. The association test P value was computed using a likelihood ratio test. Results for the X chromosome were computed similarly, with male genotypes coded as if they were homozygous diploid for the observed allele. In order to avoid issues relating to machine precision, association P-values were truncated to a minimum value of 2.2×10-308, prior to genomic inflation correction.

A principal component analysis was performed independently for each ancestry, using ∼65,000 high quality genotyped variants present in all five genotyping platforms. The analysis was performed on a subset of participants randomly sampled across all the genotyping platforms (137K, 102K, 1000K, 360K and 32K participants were used for African-American, East-Asian, European, Latino, and South-Asian, respectively). PC scores for participants not included in the analysis were obtained by projection, combining the eigenvectors of the analysis and the SNP weights.

The full GWAS summary statistics for the 23andMe discovery data set will be made available through 23andMe to qualified researchers under an agreement with 23andMe that protects the privacy of the 23andMe participants. Please visit https://research.23andme.com/collaborate/#dataset-access/ for more information and to apply to access the data.

### Power of GWAS to find associations in simulated monogenic rare diseases

We simulated monogenic rare diseases using genotyped variants on the 23andMe v5 array as known causal variants in a cohort of 4,957,230 individuals of European ancestry. We simulated a variety of disease architectures and prevalences to match those seen in real diseases we analyzed. To match real diseases, we restricted our simulations to use genotyped SNPs within genes as causal. We used a simplified version of the statistical framework described by Whiffin et al.^85^ to identify candidate causal variants and simulate case-control rare disease phenotypes (Supplementary Notes Section 1). For each simulation, we then performed association tests within a region 1 Mbp away from the gene. Only imputed SNPs were tested for association and all genotyped SNPs were excluded. The association testing used the same covariates (except genotyping platform) in the analysis and quality control as in the GWAS of rare diseases. P-values were not adjusted for inflation. To allow exploring a variety of disease architectures, we included any disease with at least 5 cases in the analysis. Power was calculated as the proportion of simulations in which the association analysis identified at least one variant with p-value < 5e-8, as would be expected in a GWAS.

### Replication of rare disease associations

Genome-wide significant associations discovered from GWAS of self-reported rare diseases were considered known if the lead SNP was within 1 Mbp of a known gene for the disease. For this analysis, genes within 1 Mbp of the lead SNP were cross-referenced against the established rare disease and gene knowledge of the Online Mendelian Inheritance in Man (OMIM) and Orphanet databases (as of September 2020). Table 1 reports hits where genes within 1 Mbp of significant lead SNPs matched known rare disease genes. For diseases with published GWASes that were not included in Orphanet or OMIM, known genes were identified through a literature search. If two nearby GWAS signals from a rare disease GWAS match the same known genes, the entries are collapsed to only include a single row in the table.

### Comparison of effect size estimates to published GWAS

To compare effect sizes from self-reported rare disease GWAS to those from published GWAS, matching disease names were identified through manual curation of the GWAS catalog (downloaded: Feb 11 2020). For a given disease, a self-reported GWAS hit and a GWAS catalog hit were considered to be the same locus if the lead SNPs for the two were in high LD (r^2^ > 0.8). When the GWAS catalog included multiple studies for a given disease, the matching of hits was performed independently for each study in the GWAS catalog.

### Replication of novel associations

We replicated the novel associations from the 23andMe discovery cohort in UKBB data. In the replication analysis, we selected the UKBB phenotypes with similar phenotypic definition as 23andMe self-reported phenotypes. We used UKBB phenotype ICD10 codes (data field: 41270) “Benign neoplasm of cranial nerves” (D33.3) in replication analysis of vestibular schwannoma. We used UKBB phenotype Non-cancer illness code (data field: 20002), self-reported, for “spontaneous pneumothorax/recurrent pneumothorax” (1126) in replication analysis of spontaneous pneumothorax. The samples in the UKBB replication analysis are restricted to British samples (based on data field 21000: 1001) with no kinship found (based on data field 22021: 0). Using this approach, 232 cases and 291,991 controls of benign neoplasm of cranial nerves and 707 cases and 291,516 controls of spontaneous pneumothorax/recurrent pneumothorax are included in the replication analysis. We applied the PHESANT^86^ software to perform a logistic regression assuming an additive model for allelic effects with adjustment for age, sex, genotype chip, assessment centre and the first 10 genetic principal components.

### Trans-ethnic replication of known associations

Known genome-wide significant associations that replicated in our European GWAS were further tested in our non-European cohorts. We selected phenotypes where any of the African American, East Asian, Latino, and South Asian cohorts had more than 30 cases. We performed association analysis separately in each population and included the same set of covariates as in the European GWAS: age, gender, genotyping platform, and 5 population-specific principal components. We first performed association tests on lead SNPs from table 1. To examine whether different SNPs were more significant in other populations, we also tested all variants within 200 kbp of the lead SNP from the European GWAS.

## Acknowledgements

We thank the research participants and employees of 23andMe for making this work possible. Some analyses in this research were conducted using the UK Biobank Resource under application number 54124, Learning models for predicting lab values from genetic data.

The following members of the 23andMe Research Team contributed to this study: Stella Aslibekyan, Adam Auton, Elizabeth Babalola, Robert K. Bell, Nic Berns, Jessica Bielenberg, Katarzyna Bryc, Emily Bullis, Daniella Coker, Gabriel Cuellar Partida, Devika Dhamija, Sayantan Das, Sarah L. Elson, Teresa Filshtein, Kipper Fletez-Brant, Pierre Fontanillas, Will Freyman, Pooja M. Gandhi, Karl Heilbron, Barry Hicks, David A. Hinds, Ethan M. Jewett, Yunxuan Jiang, Katelyn Kukar, Keng-Han Lin, Maya Lowe, Jey McCreight, Matthew H. McIntyre, Steven J. Micheletti, Meghan E. Moreno, Joanna L. Mountain, Priyanka Nandakumar, Elizabeth S. Noblin, Jared O’Connell, Aaron A. Petrakovitz, G. David Poznik, Morgan Schumacher, Anjali J. Shastri, Janie F. Shelton, Jingchunzi Shi, Suyash Shringarpure, Vinh Tran, Joyce Y. Tung, Xin Wang, Wei Wang, Catherine H. Weldon, Peter Wilton, Alejandro Hernandez, Corinna Wong, Christophe Toukam Tchakouté.

