## Supplementary Text and Figures for "Large-scale trans-ethnic replication and discovery of genetic associations for rare diseases with self-reported medical data"

#### Supplementary Notes

**Section 1: Simulation of rare diseases**

To assess the power of GWAS in simulated data, we simulated monogenic autosomal dominant and recessive rare diseases using genotyped variants on the 23andMe v5 array as known causal variants.
We simulated diseases with a range of penetrances, numbers of causal variants and prevalences within the European population. To match real diseases, we restricted our simulations to genotyped SNPs within genes. We assumed causal variants were independent of each other in choosing sets of causal variants. For each parameter setting, we simulated 40 disease phenotypes.

**Simulation of dominant disease:**

For a given combination of prevalence, penetrance, number of causal variants, and gene, we simulated a dominant disease phenotype as follows:

- Under the dominant model, for a given set of causal variants, prevalence is equal to penetrance times twice the sum of the frequencies of the causal alleles. Therefore, twice the desired sum of causal allele frequencies was set equal to prevalence/penetrance.
- We sampled the desired number of causal variants from genotyped SNPs within the gene and computed the sum of frequencies of minor allele carriers
- The sampled set of causal variants was accepted if twice the sum of frequencies of minor allele carriers was within 20% of the desired prevalence/penetrance, else the sampling from the previous step was repeated (at most 400 times).
- Using the selected set of causal variants, individuals were identified as candidate cases if they had a minor allele for any of designated causal variants, and as controls otherwise.
- Among candidate cases, individuals were classified as cases with probability equal to the penetrance, and as controls otherwise.

**Simulation of recessive disease:**

For a given combination of prevalence, penetrance, number of causal variants, and gene, we simulated a recessive disease phenotype as described below. Our model considers only homozygous carriers of causal variant minor alleles as cases. Compound heterozygous individuals are considered as controls.

- Under the recessive model, for a given set of causal variants, prevalence is equal to penetrance times the sum of the squared frequencies of the causal alleles. Therefore, the desired sum of squared causal allele frequencies was set equal to prevalence/penetrance.
- We sampled the desired number of causal variants from genotyped SNPs within the gene and computed the sum of squares of frequencies of minor allele carriers
- The sampled set of causal variants was accepted if the sum of frequencies of minor allele carriers was within 20% of the desired prevalence/penetrance, else the sampling from the previous step was repeated (at most 400 times).
- Using the selected set of causal variants, individuals were identified as candidate cases if they had two copies of the minor allele for any of designated causal variants, and as controls otherwise.
- Among candidate cases, individuals were classified as cases with probability equal to the penetrance, and as controls otherwise.

For each simulation, we then performed association tests within a region of 1 Mbp on either side of the gene. Only imputed SNPs were tested for association and all genotyped SNPs were excluded. The association testing used the same covariates in the analysis and quality control as in the GWAS of real diseases, except for genotyping platform covariates since all individuals were genotyped on the same platform. Similar to the original GWAS, controls were downsampled to only include 5% of all controls. P-values were not adjusted for inflation.

**Section 2: Trait ascertainment**

Subjects with rare diseases were identified through self-report in web-based surveys. Subject responses were combined from their answers to the following questions:

1. Health history survey (2015 onwards): This is a general medical history survey which asks research participants about their medical conditions through a question format such as “Have you ever been diagnosed with, or treated for <condition name>?
   Some conditions, which are better understood as subtypes of other conditions, or as a cause of other conditions, were asked as follow up questions. For example, to ascertain Factor V Leiden, the following set of questions were used in sequence:

   Have you ever been diagnosed with, or treated for, a blood clot or condition causing repeated blood clots? <Participants selecting “yes” received the next followup question>
   Did your doctor tell you that your blood clotting condition was mainly caused by any of the following? <Participants selecting hypercoagulation received the next followup question>
   Which blood clotting condition were you diagnosed with most recently? <Participants selecting Factor V Leiden ascertained as cases>

2. Health history survey free text (2015 onwards): Research participants are able to report on medical conditions not covered by the general question format in #1 through a free-text question towards the end of the survey, "Have you ever been diagnosed with, or treated for, any other condition not yet mentioned?". Responses to this question were parsed using text mining to extract reports of rare conditions of interest.
3. Rare disease survey (2017 onwards): This is a web-based survey with a special focus on rare diseases, where participants can report on the 9000+ disease terms through an autocomplete feature. Participants are asked to enter at least the first three letters of the rare condition(s) they have been diagnosed with, and a list of matching entries appears from which participants can select their response. The complete dictionary for these rare disease terms were derived from the Orphanet database (http://www.orphadata.org/cgi-bin/rare_free.html). If a participant is unable to find their condition in the dropdown list, they can select "Other" and report their response through a free-text followup question.
4. Disease specific surveys: In addition to the health history survey and rare disease survey, 23andMe employs many disease specific surveys to gather additional deep dive information about conditions such as cancer, arthritis, multiple sclerosis. Case ascertainment for certain rare conditions (e.g. rare cancers) was made through one of these surveys.
5. Quick questions: These are stand-alone questions which ask participants about specific diagnoses. The standard format is similar to the health history survey question "Have you ever been diagnosed with, or treated for, <condition name>"

Sources 1,3,4 and 5 were combined by defining cases as the union of cases among these sources for each participant; the controls were defined as individuals who met the criteria for controls for either of the above conditions (1,3,4 or 5) and were not defined as cases for either condition (1,3,4 or 5). We then incorporated the combined responses above to source 2 and the free text data from source 3, by keeping the first non-missing response among these sources for each participant, evaluated in the specified order where the combined responses were evaluated first.

A summary of all data sources for each of the conditions in Table 1 and 2 are presented in Table S3.

### Supplementary Figures

a

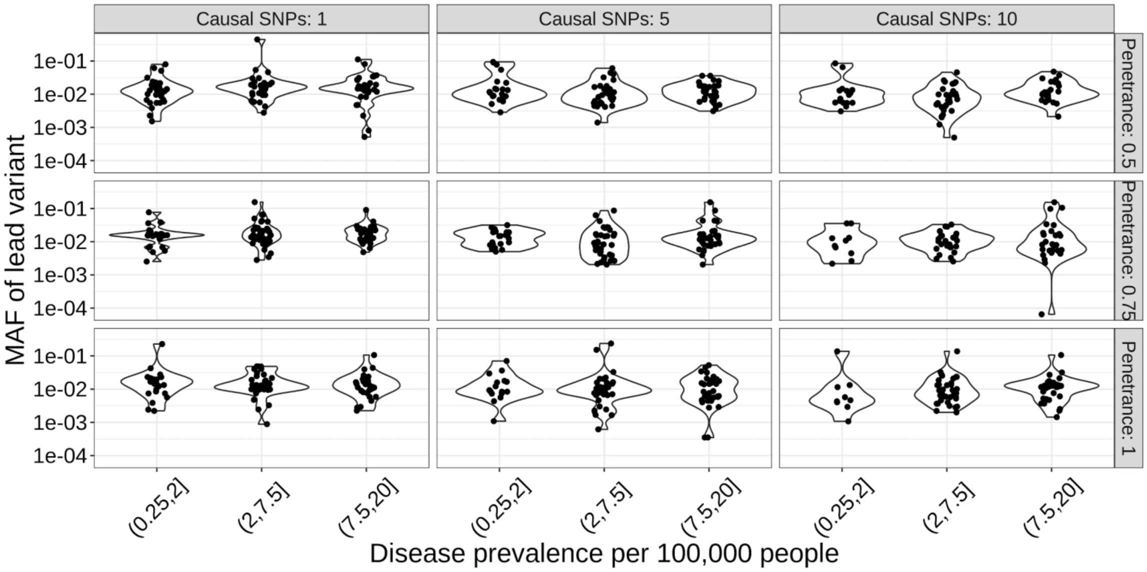


b

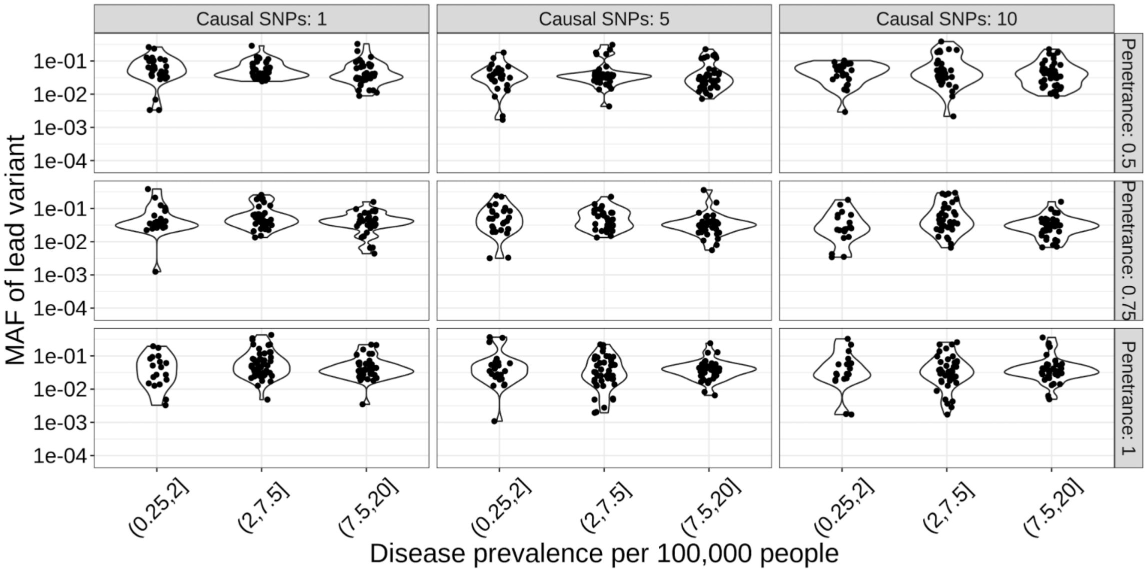


Supplementary Figure 1: Minor allele frequency of lead SNP in simulations where GWAS of rare disease finds a genome-wide significant hit near the simulated causal gene under a (a) dominant inheritance model (b) recessive inheritance model.

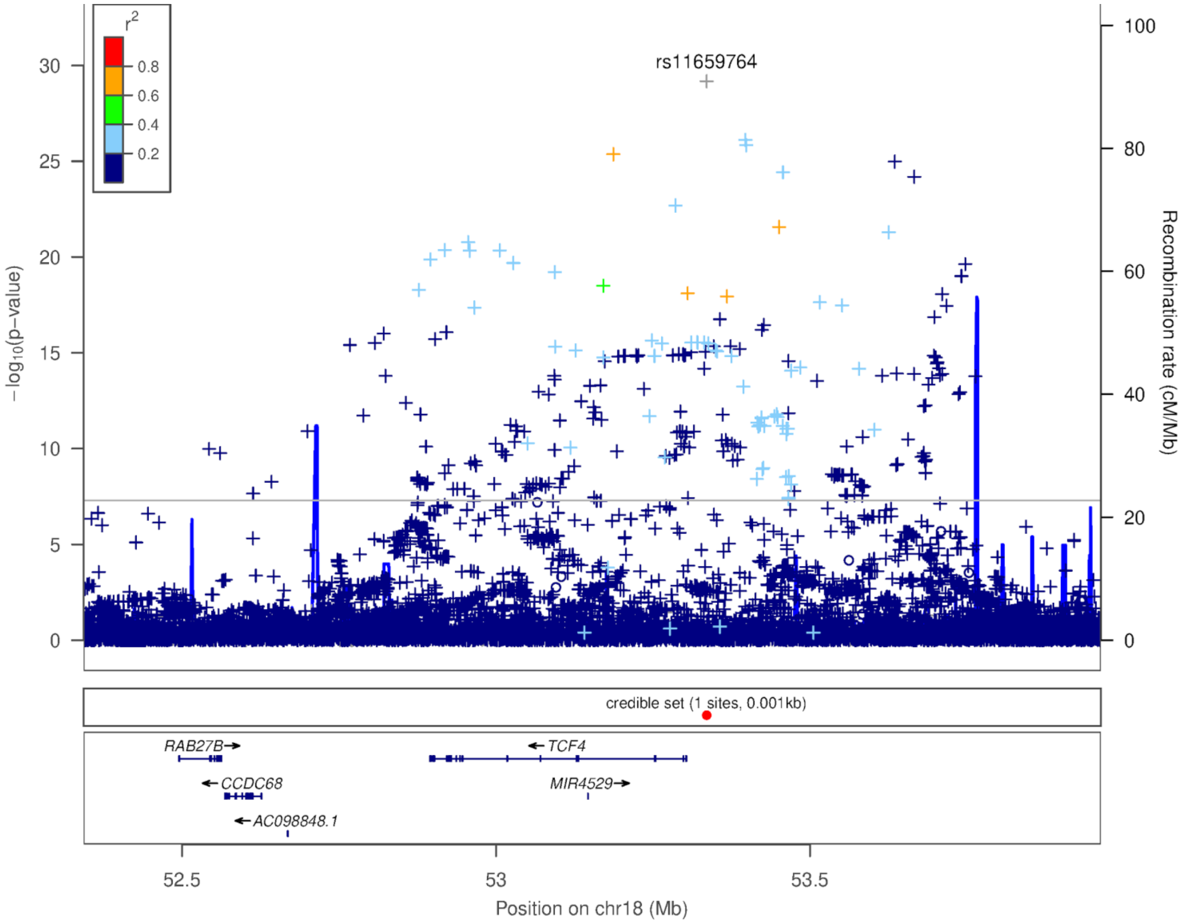


Supplementary Figure 2: Locuszoom plot for Fuch's corneal dystrophy

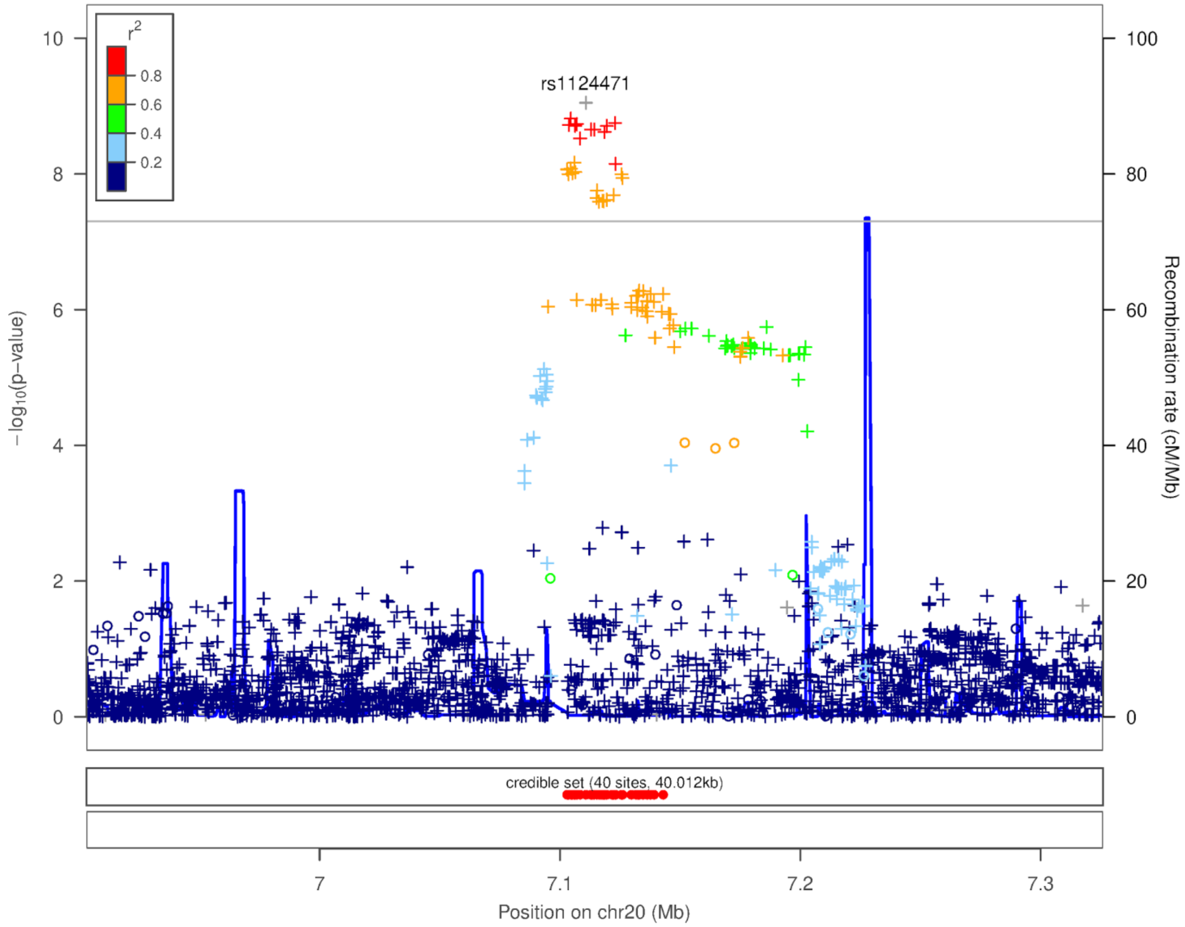


Supplementary Figure 3: Locuszoom plot for craniosynostosis chromosome 20 association

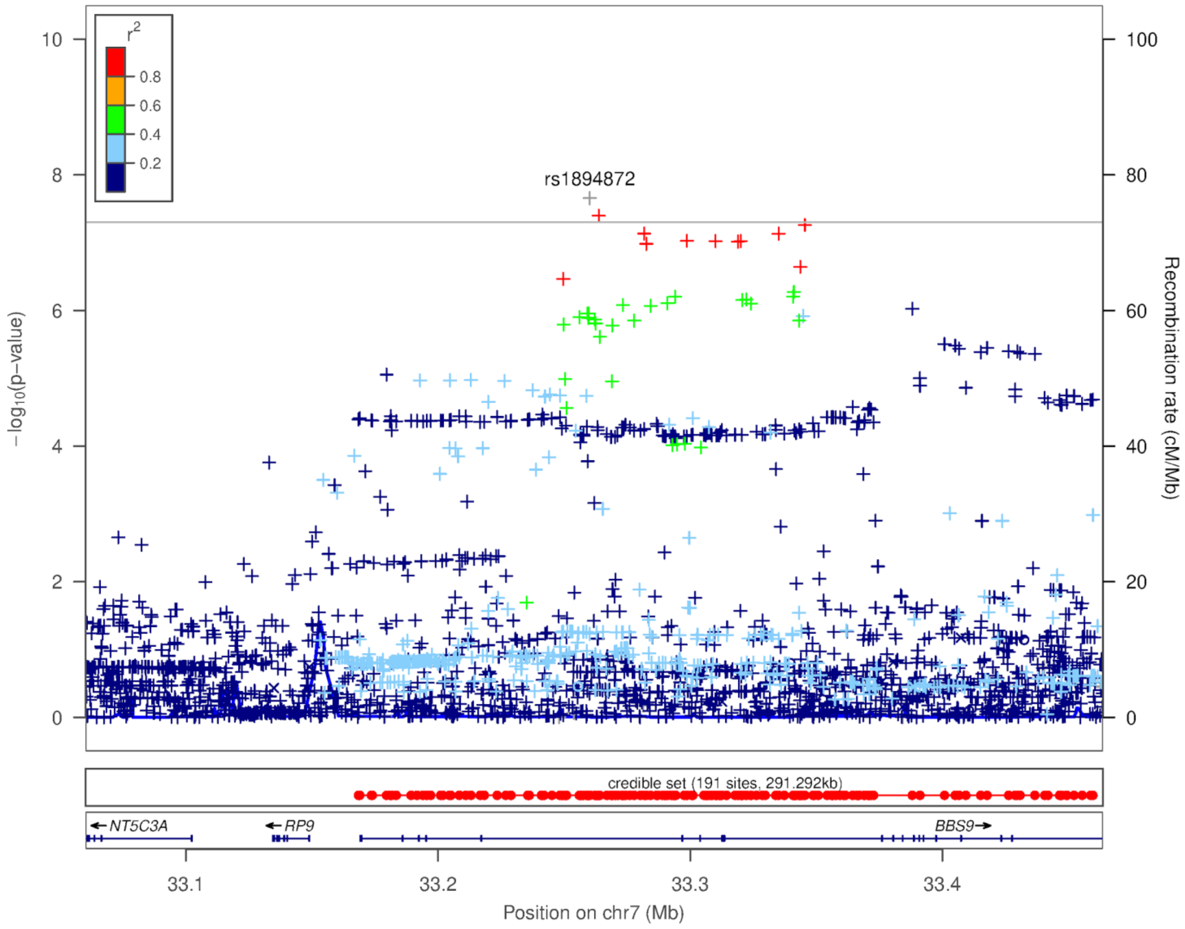


Supplementary Figure 4: Locuszoom plot for craniosynostosis chromosome 7 association

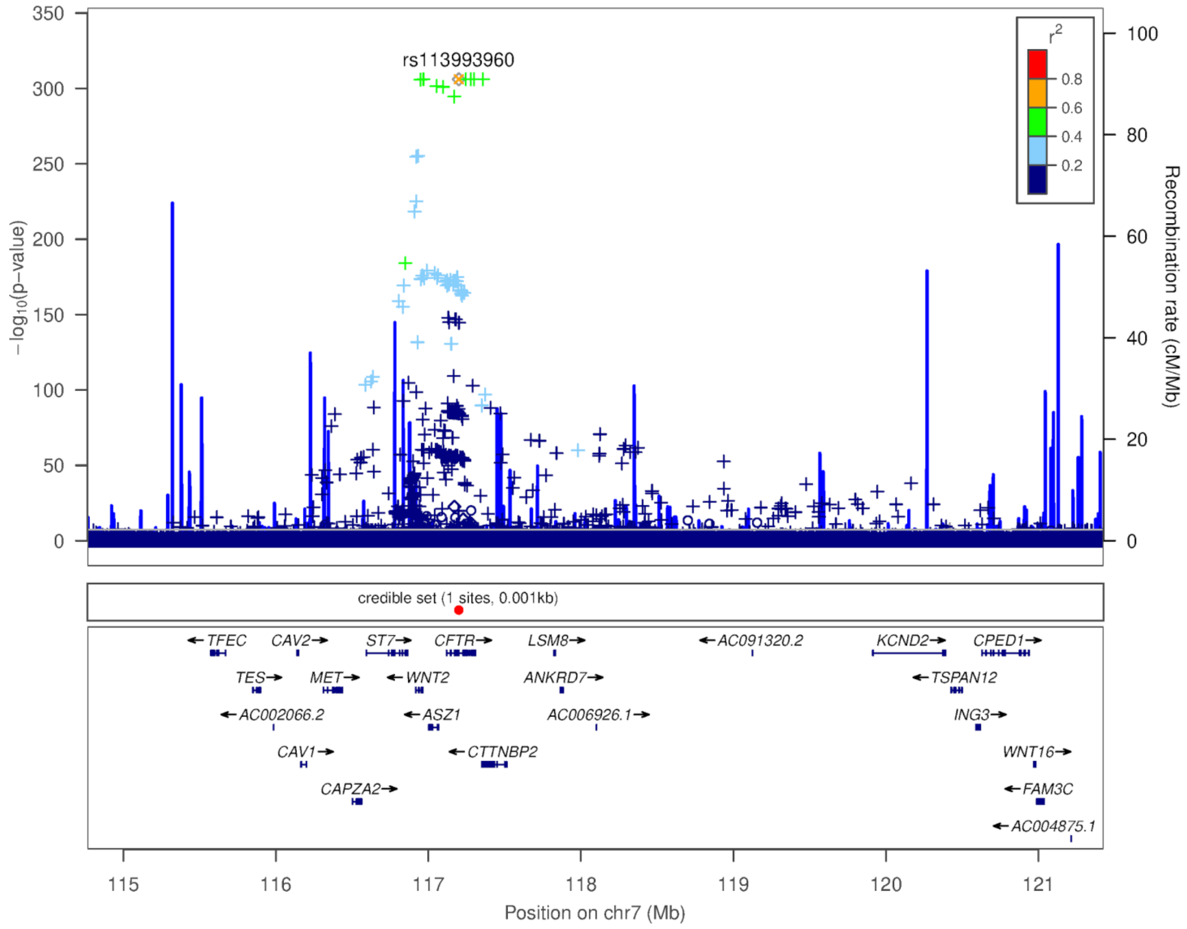


Supplementary Figure 5: Locuszoom plot for cystic fibrosis association

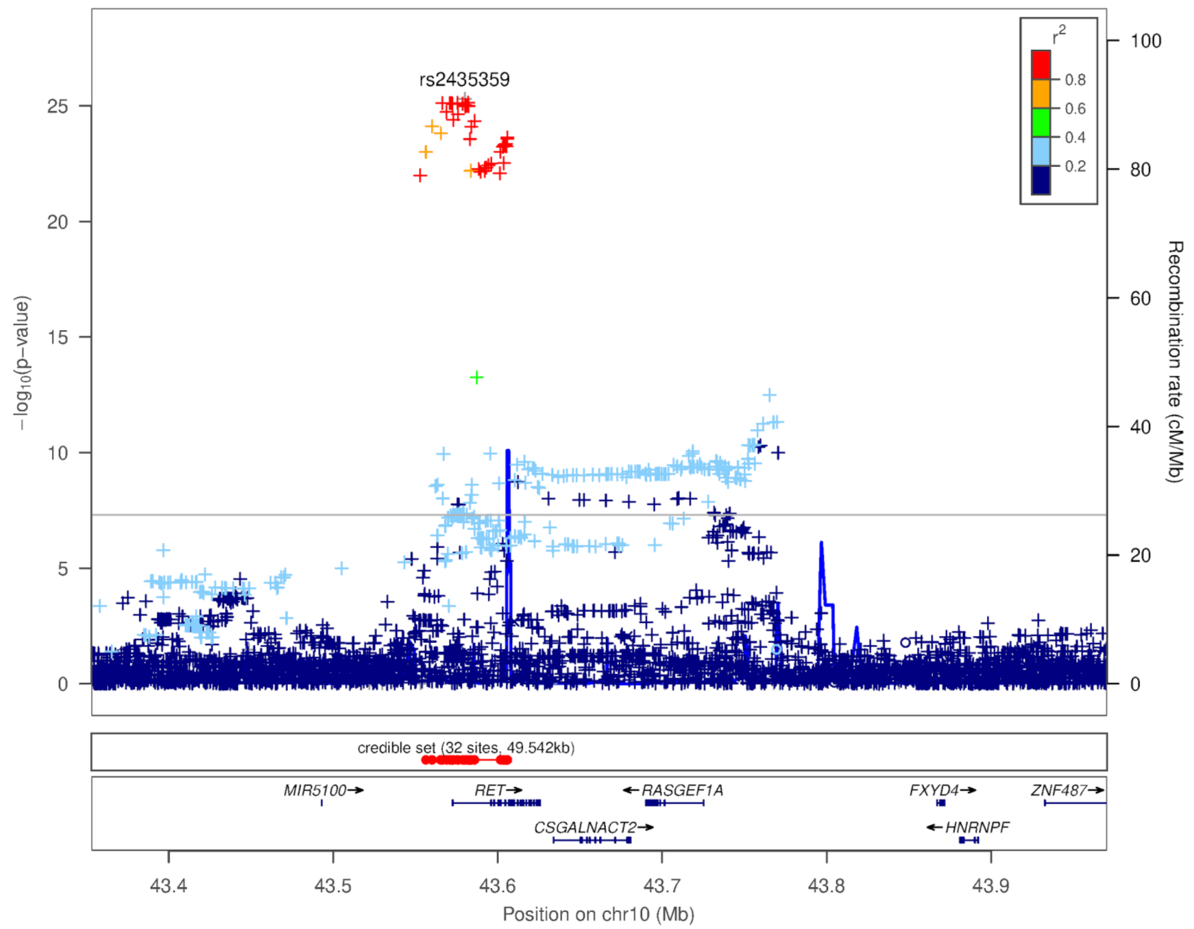


Supplementary Figure 6: Locuszoom plot for hirschsprung disease association

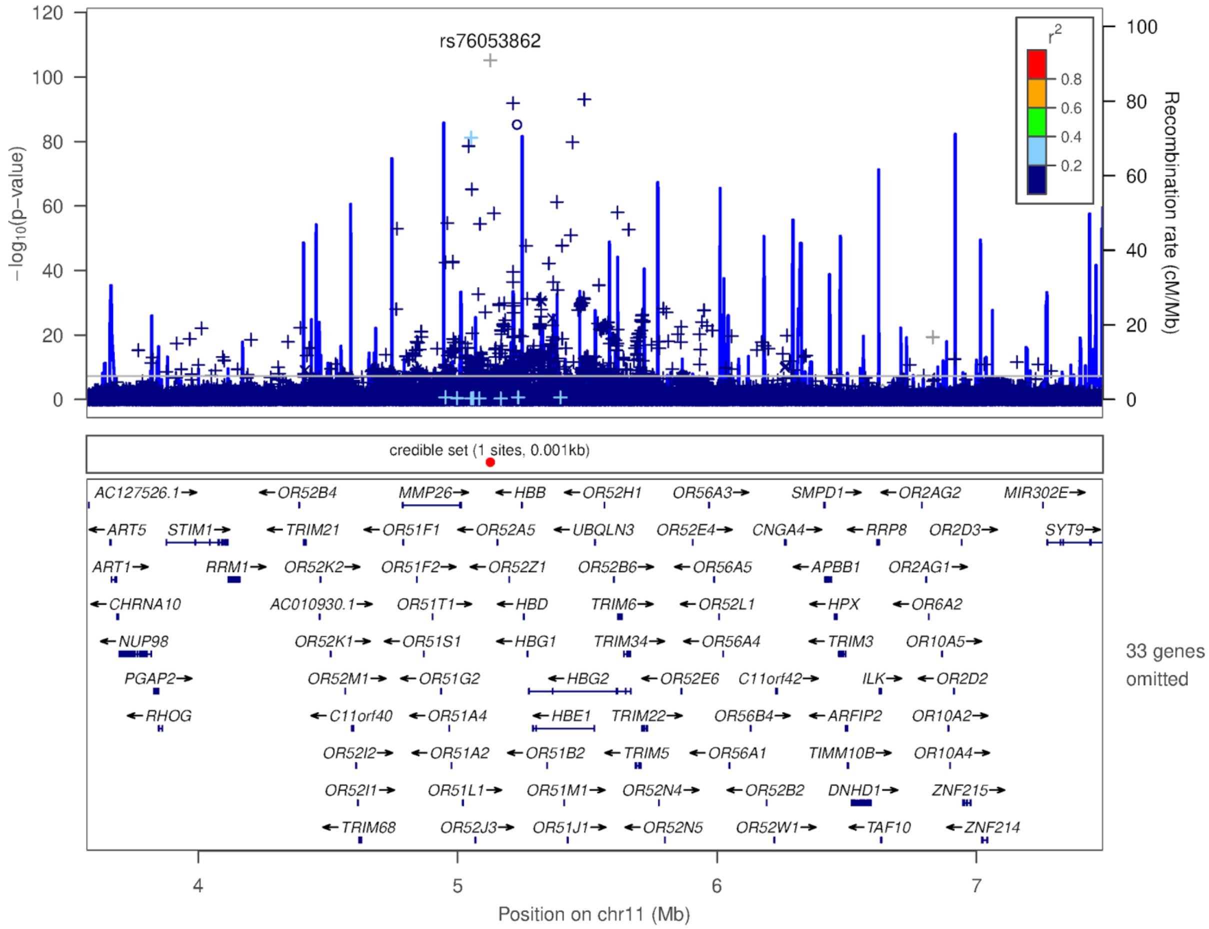


Supplementary Figure 7: Locuszoom plot for beta thalassemia association

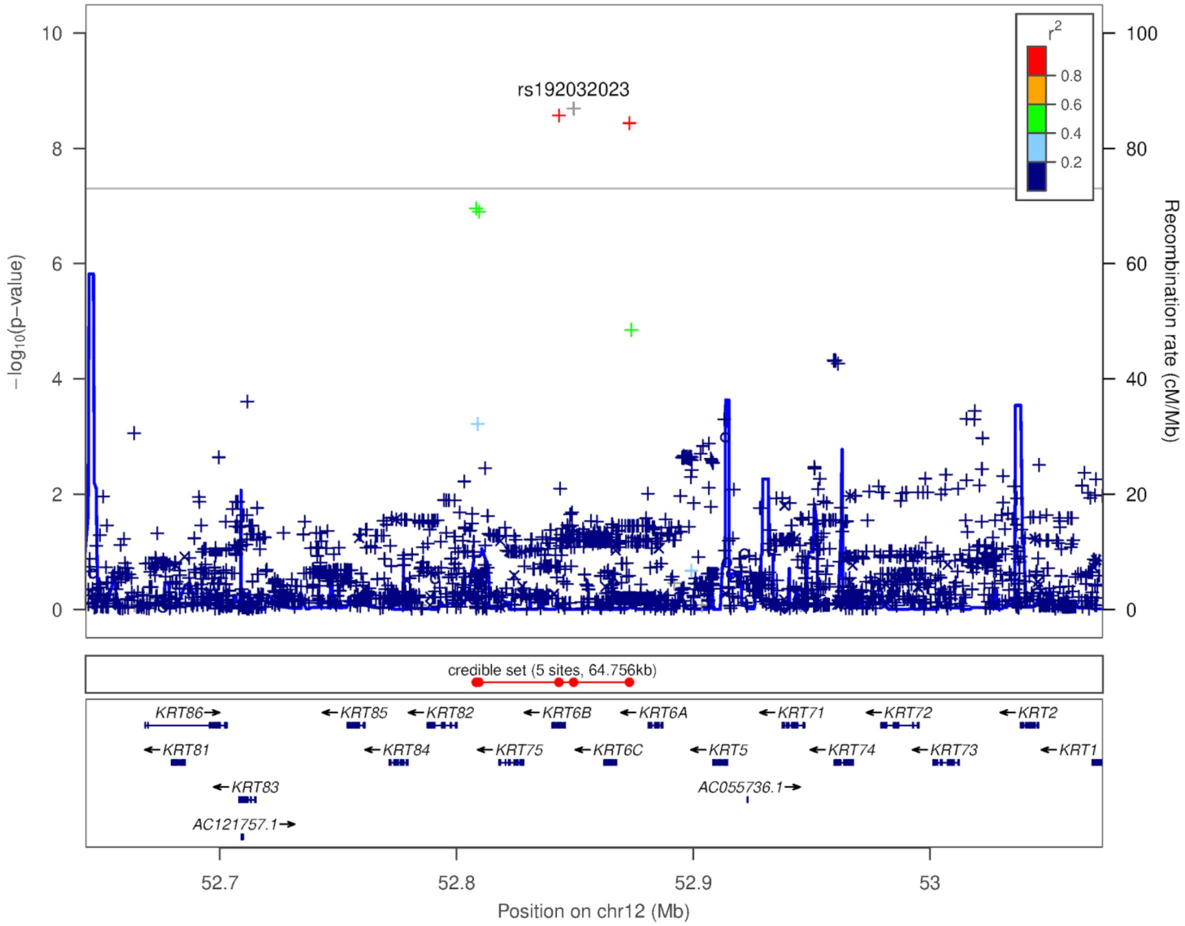


Supplementary Figure 8: Locuszoom plot for epidermolysis bullsoa simplex association

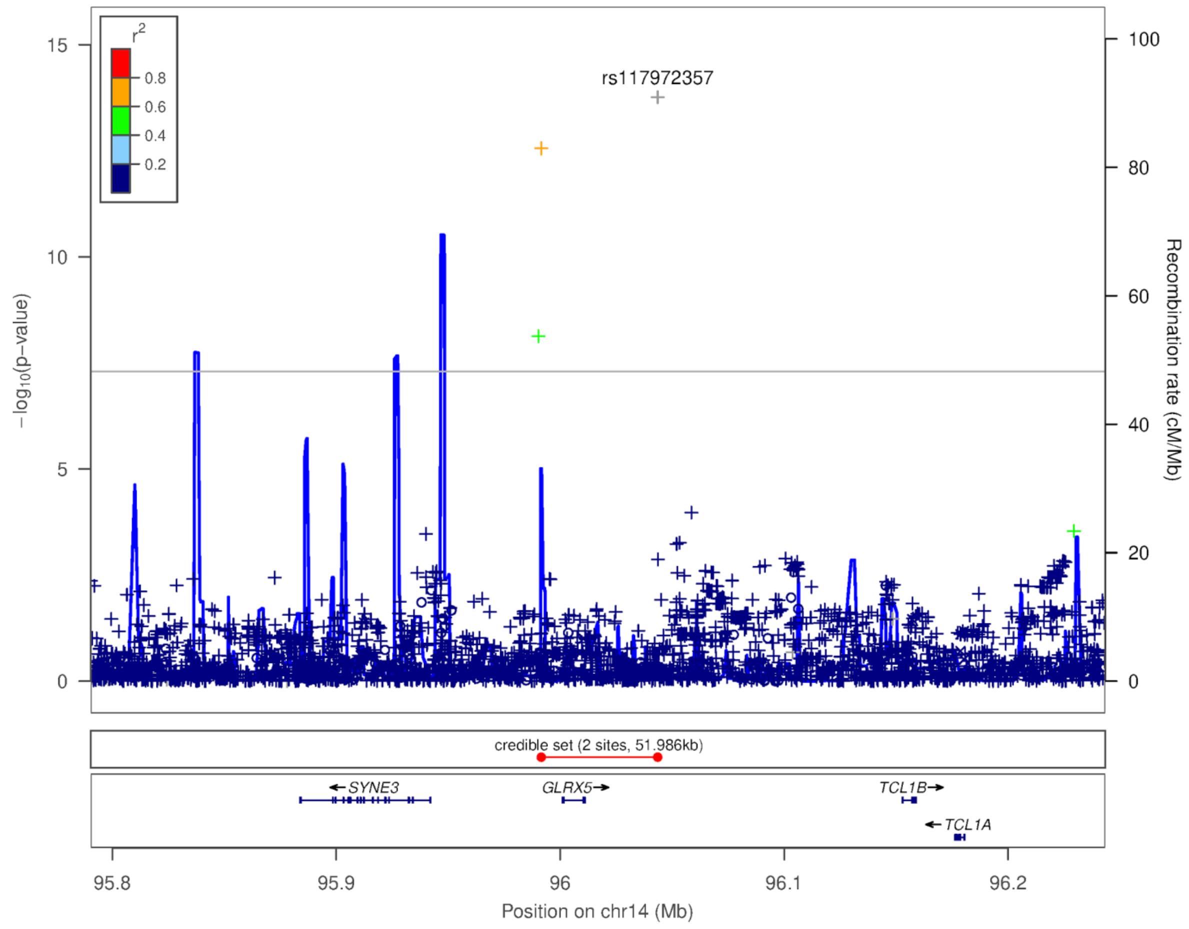


Supplementary Figure 9: Locuszoom plot for waldenstrom macrogloblulinemia chromosome 14 association

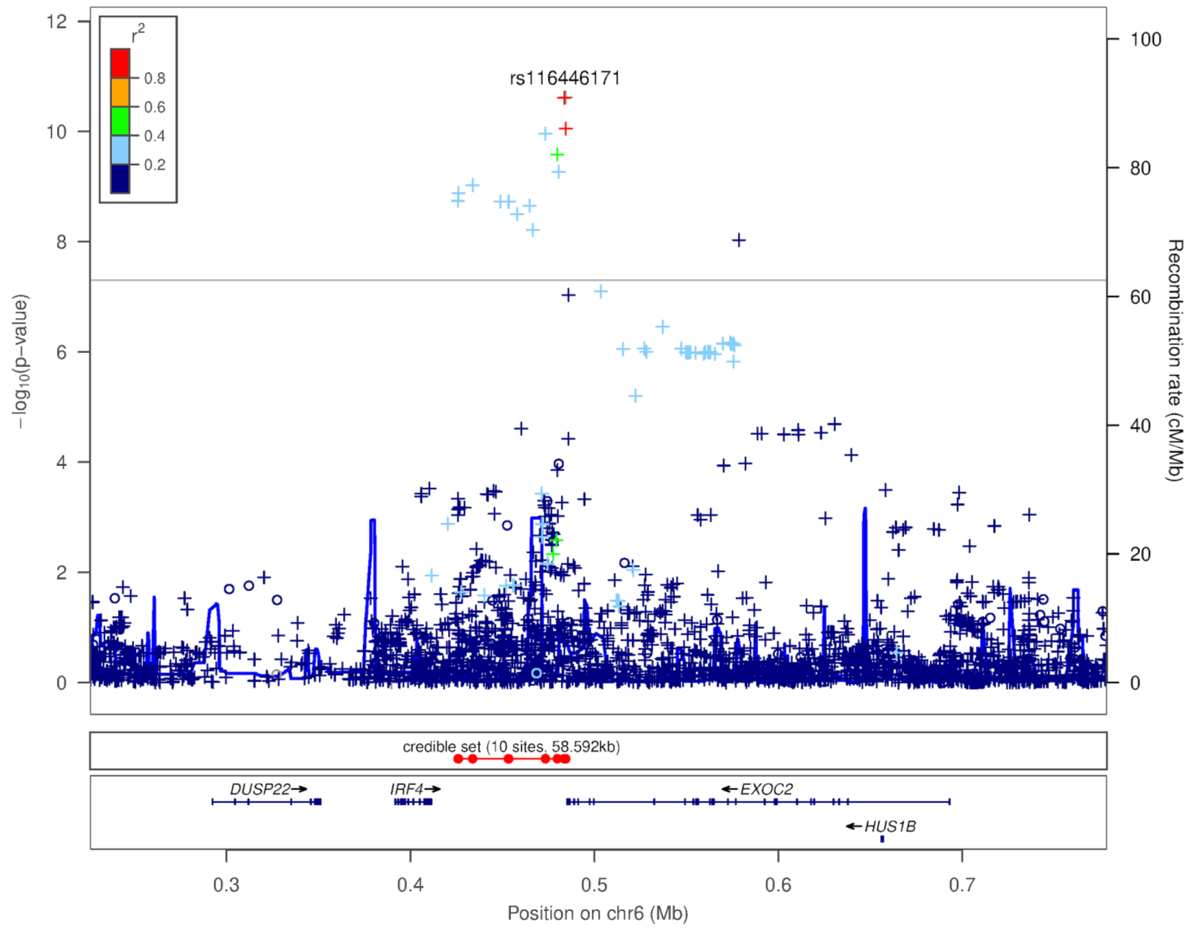


Supplementary Figure 10: Locuszoom plot for waldenstrom macroglobulinemia chromosome 6 association

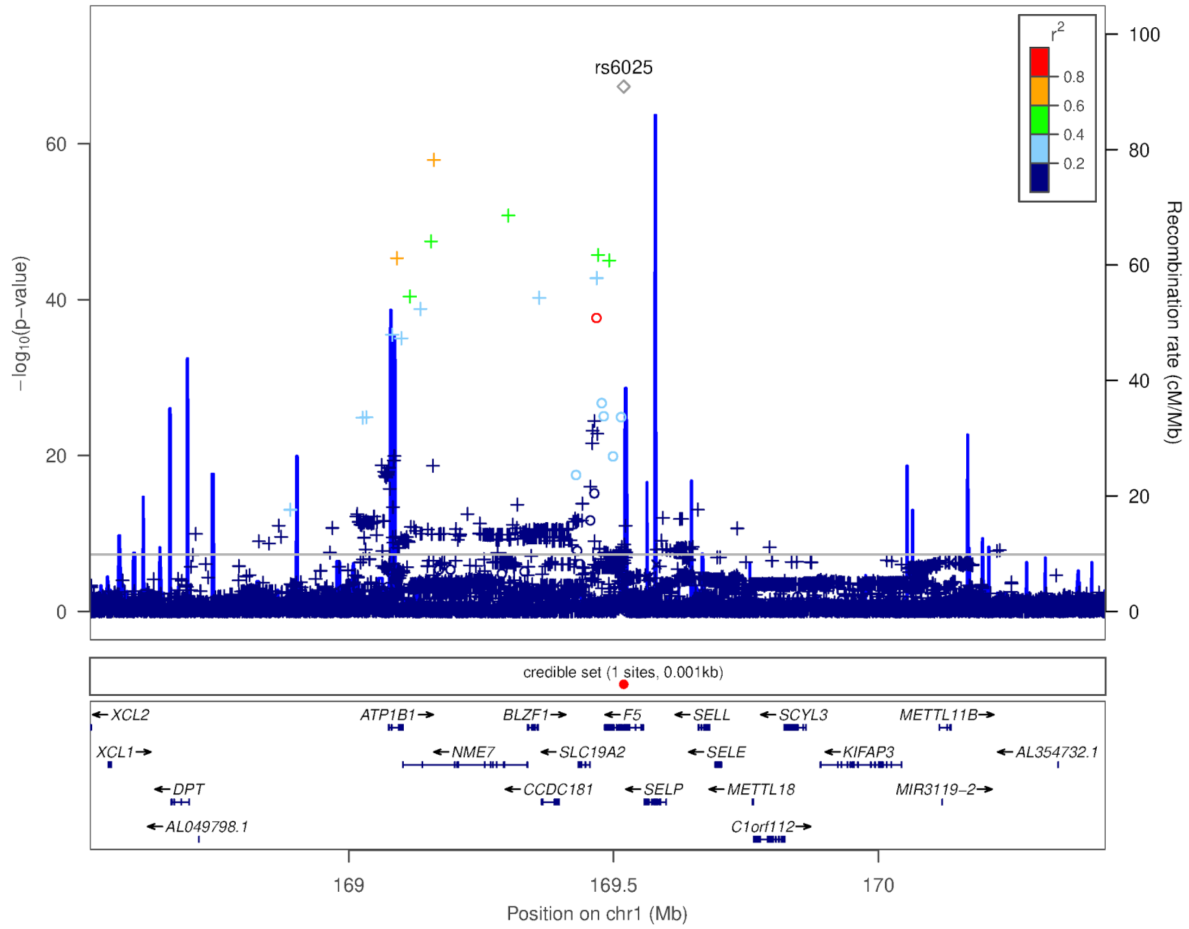


Supplementary Figure 11: Locuszoom plot for Factor V deficiency association

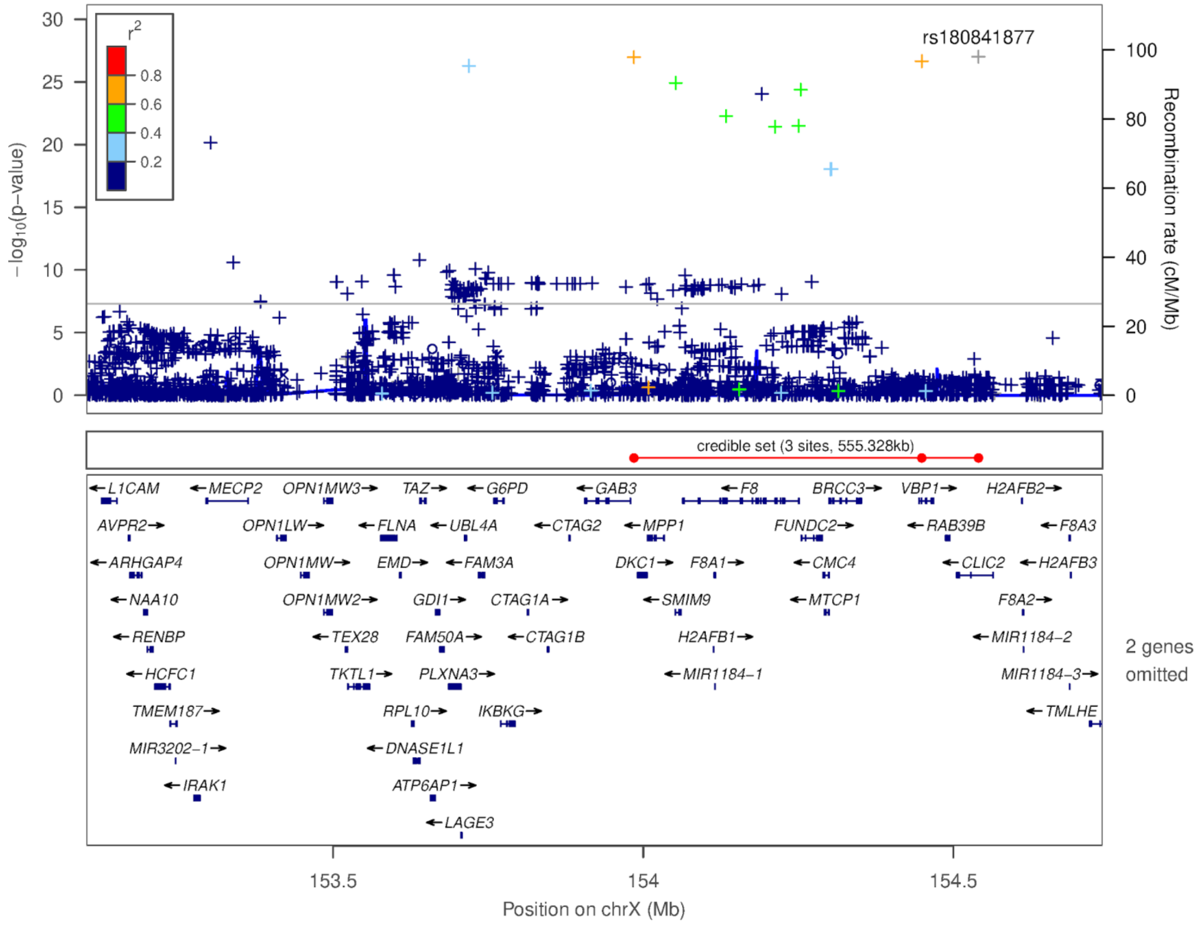


Supplementary Figure 12: Locuszoom plot for glucose-6-phosphate dehydrogenase deficiency association

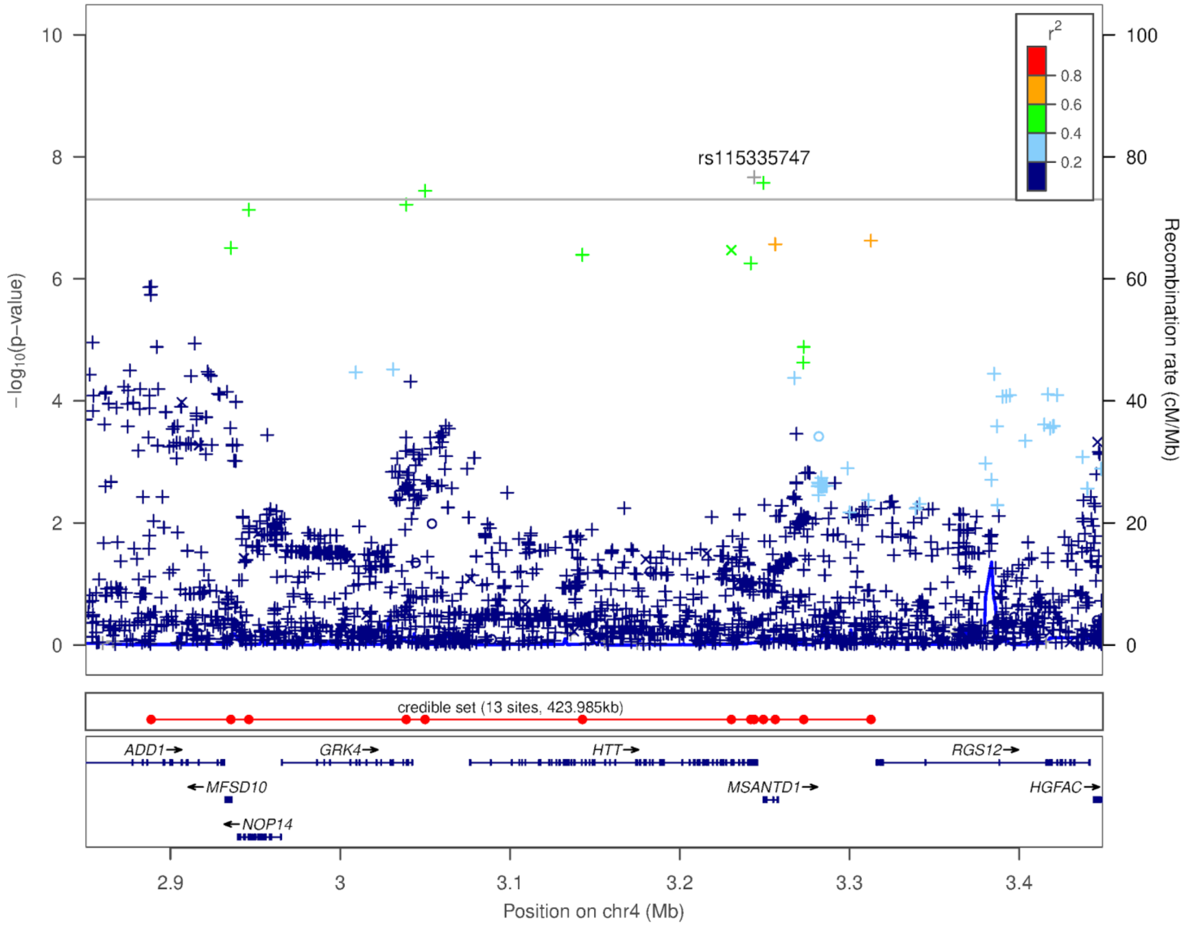


Supplementary Figure 13: Locuszoom plot for Huntington's disease association

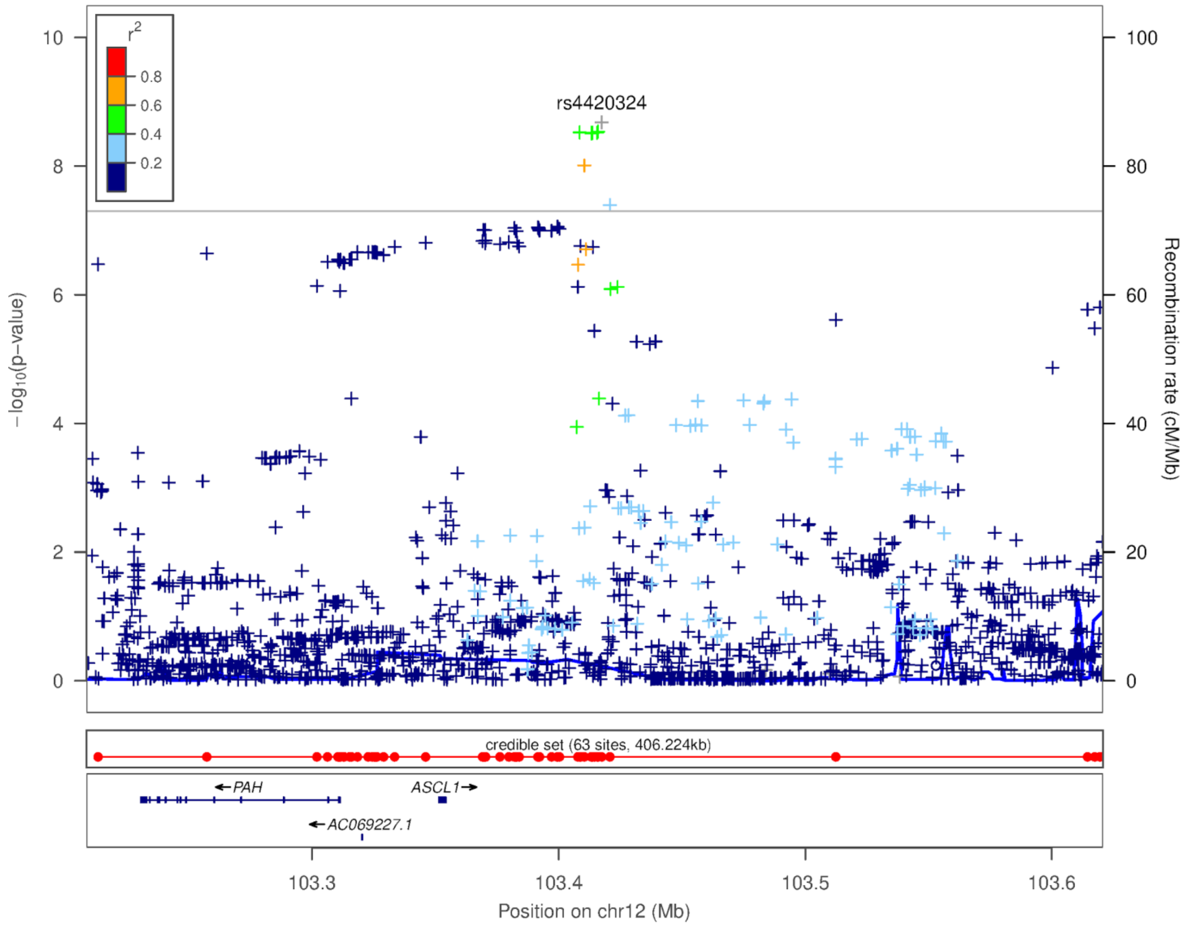


Supplementary Figure 14: Locuszoom plot for phenylketonuria association

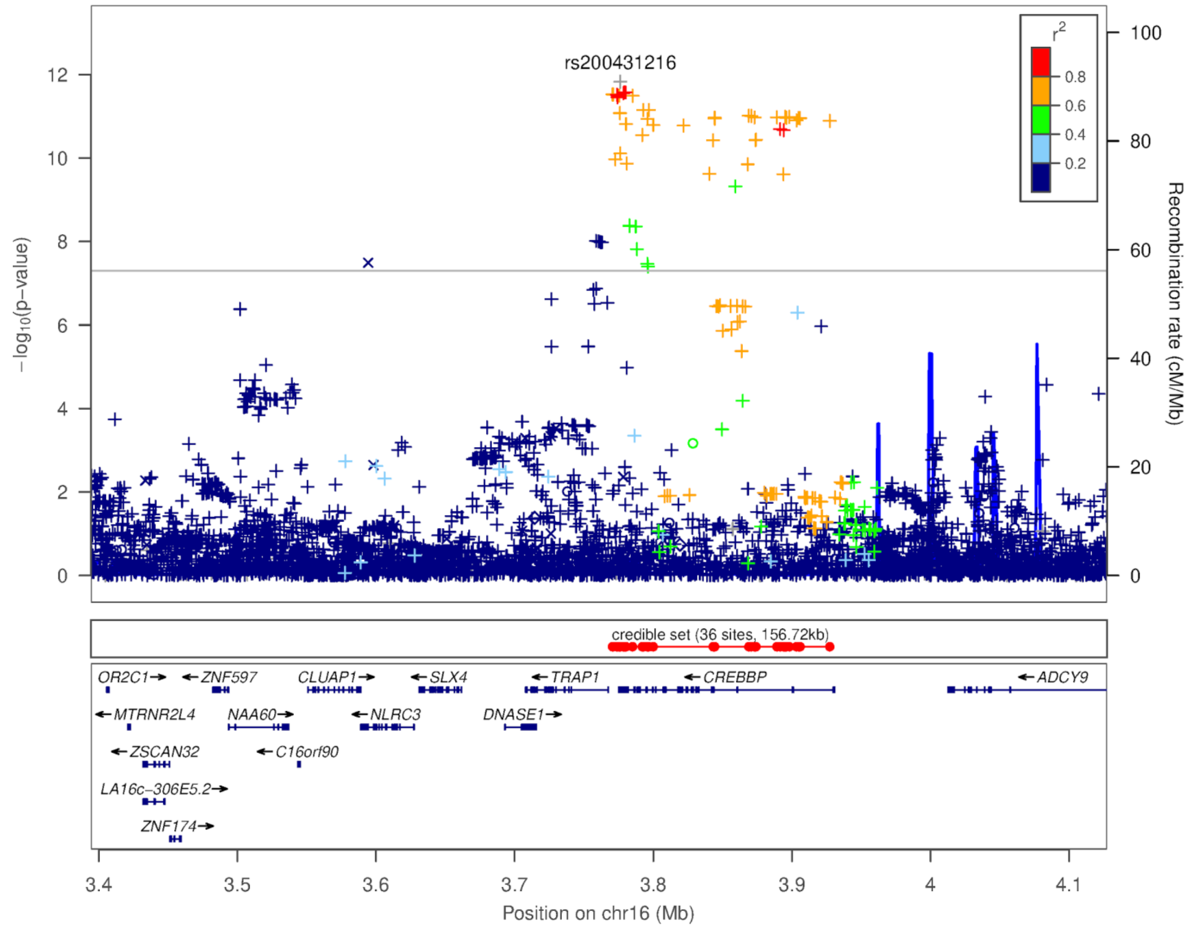


Supplementary Figure 15: Locuszoom plot for familial mediterranean fever association

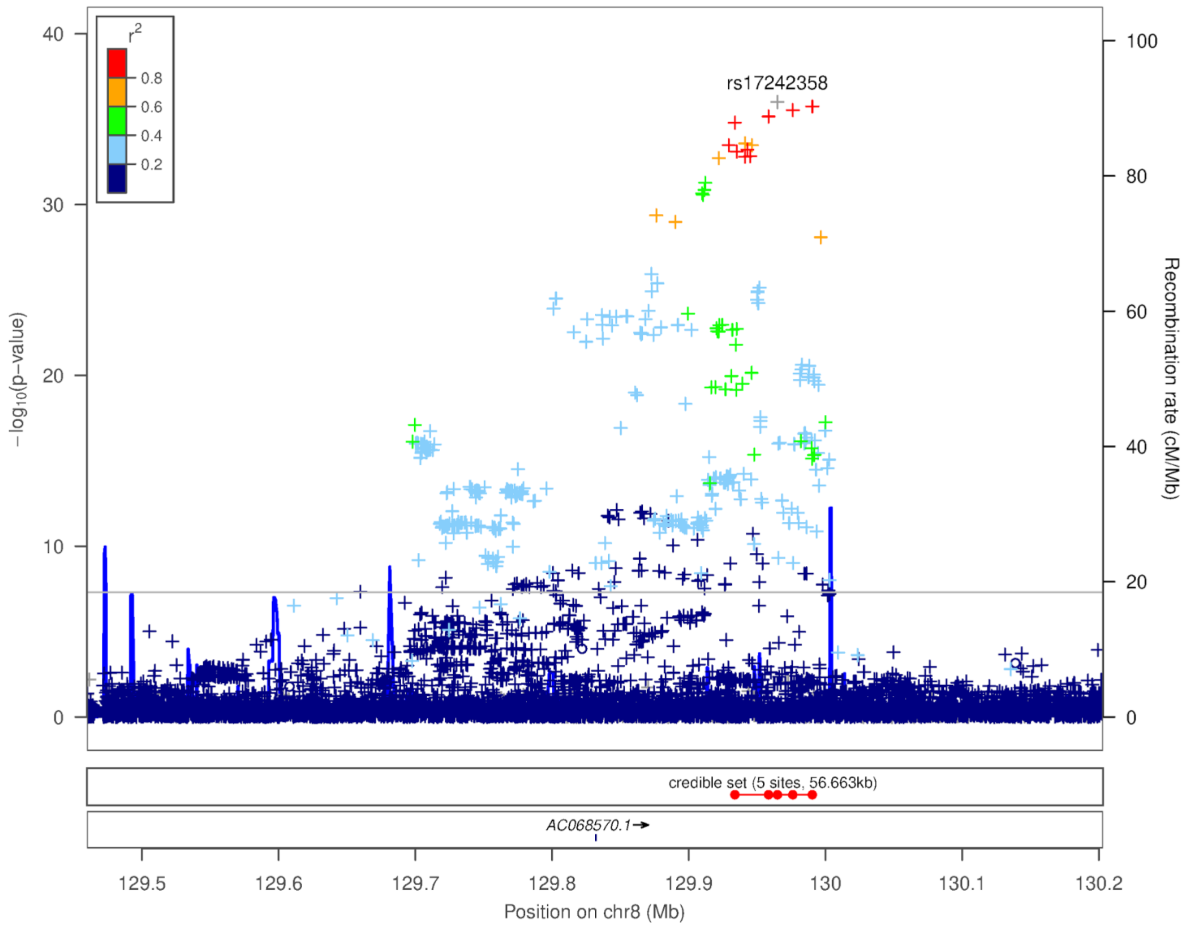


Supplementary Figure 16: Locuszoom plot for cleft lip chromosome 8 association

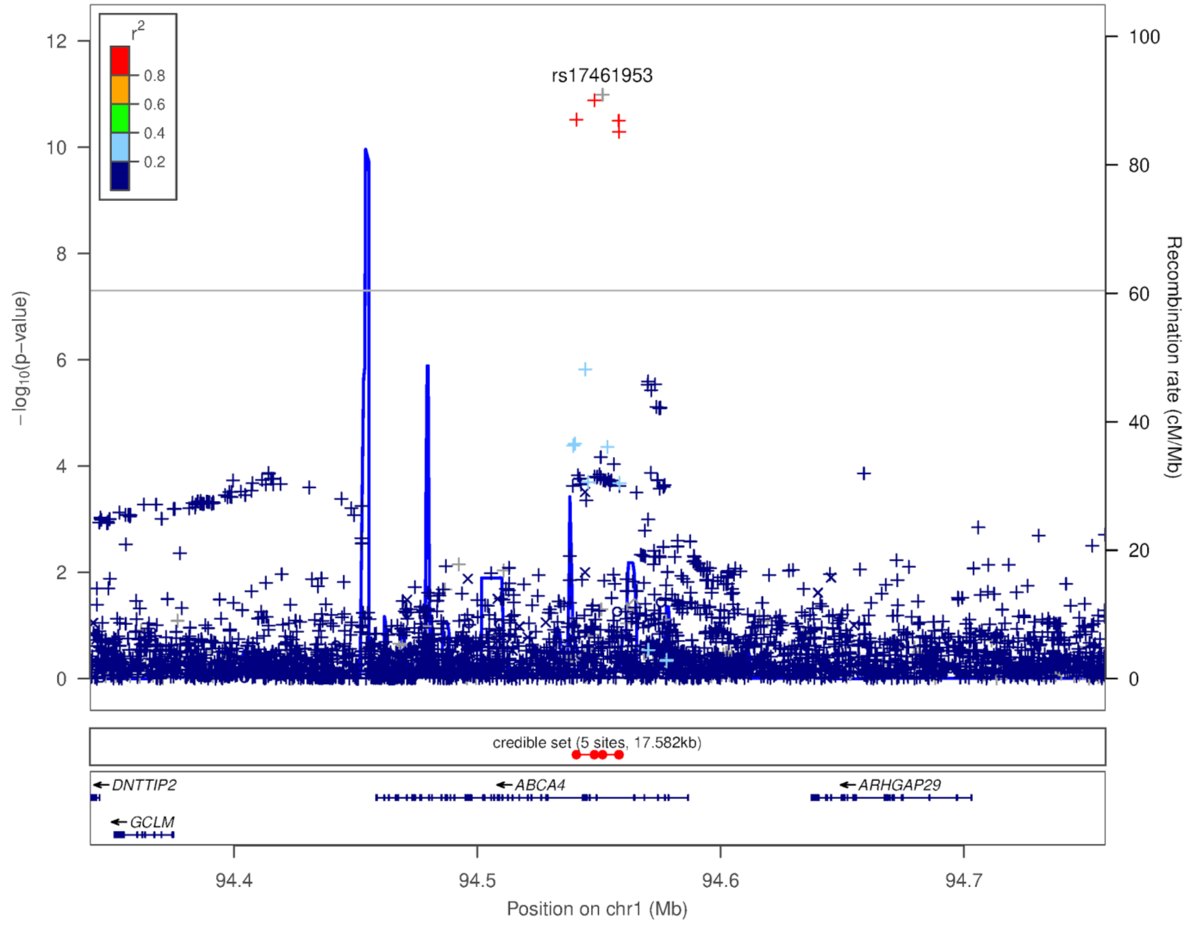


Supplementary Figure 17: Locuszoom plot for cleft lip chromosome 1p22.1 association

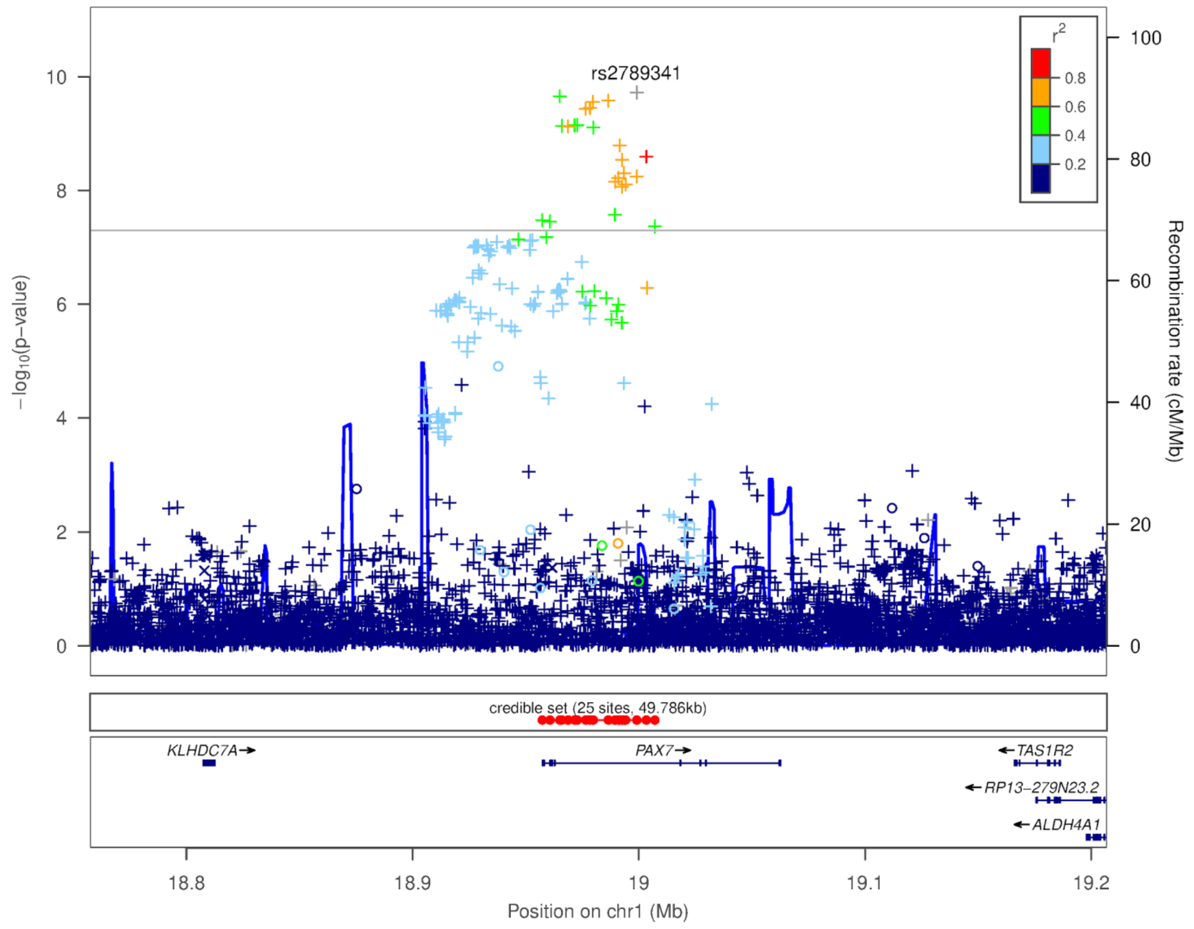


Supplementary Figure 18: Locuszoom plot for cleft lip chromosome 1p36.13 association

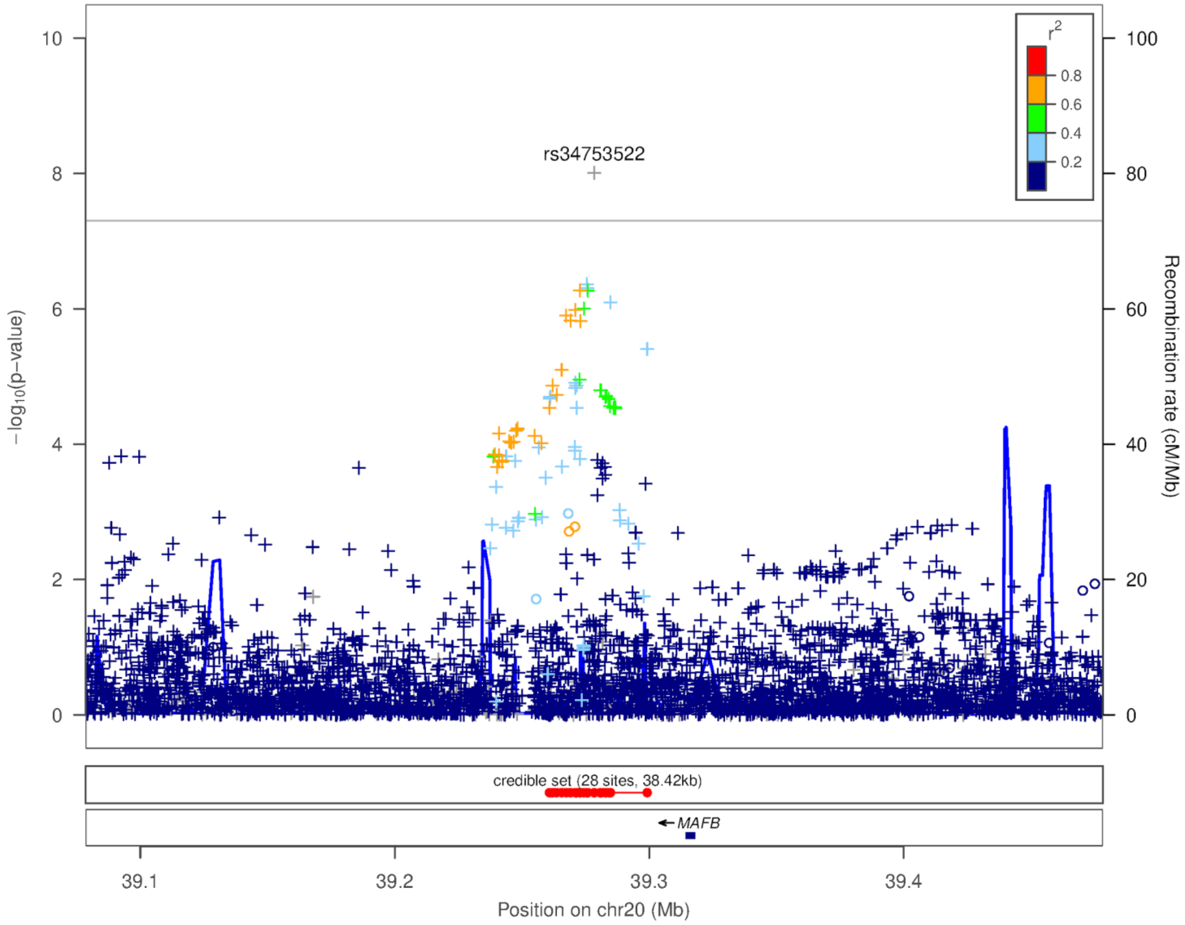


Supplementary Figure 19: Locuszoom plot for cleft lip chromosome 20 association

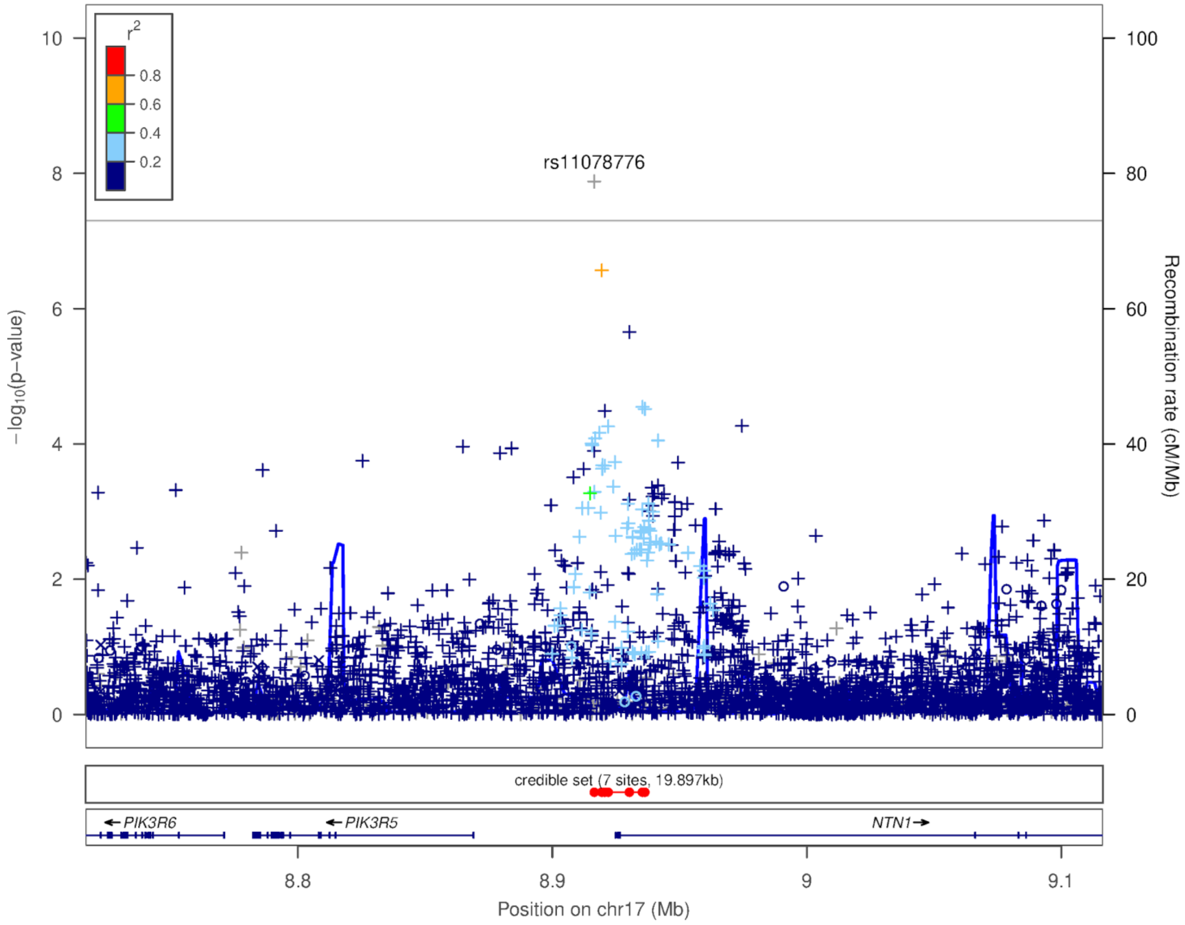


Supplementary Figure 20: Locuszoom plot for cleft lip chromosome 17 association

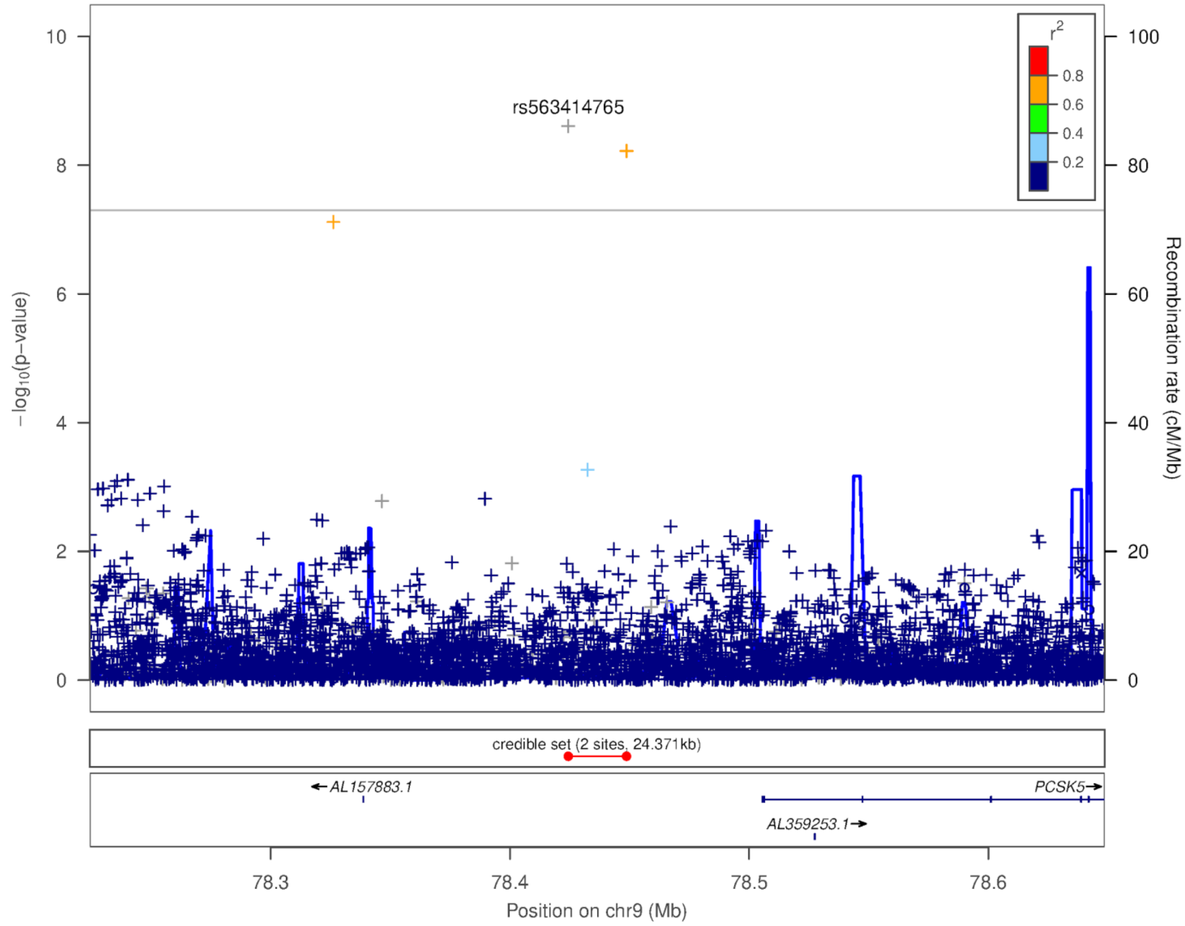


Supplementary Figure 21: Locuszoom plot for cleft lip chromosome 9 association

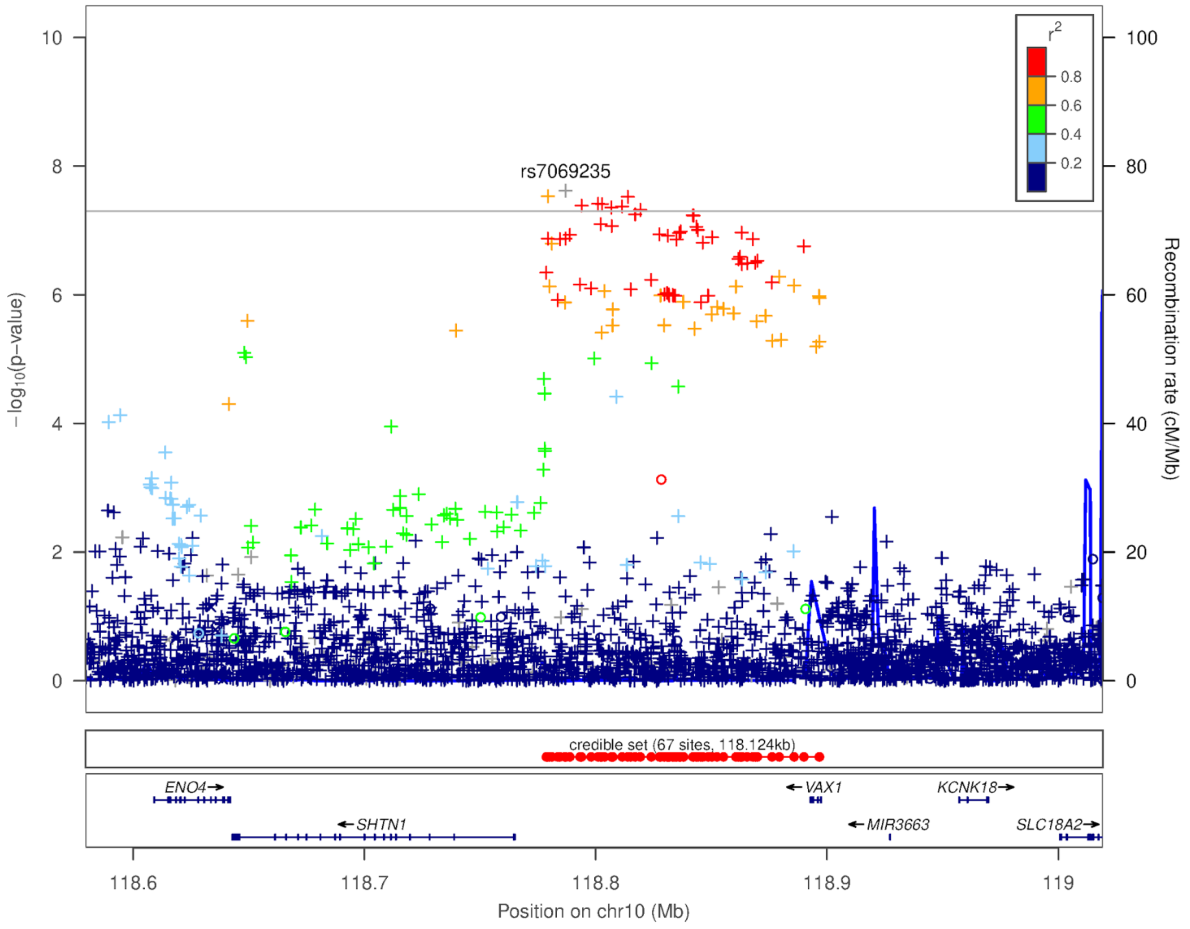


Supplementary Figure 22: Locuszoom plot for cleft lip chromosome 10 association

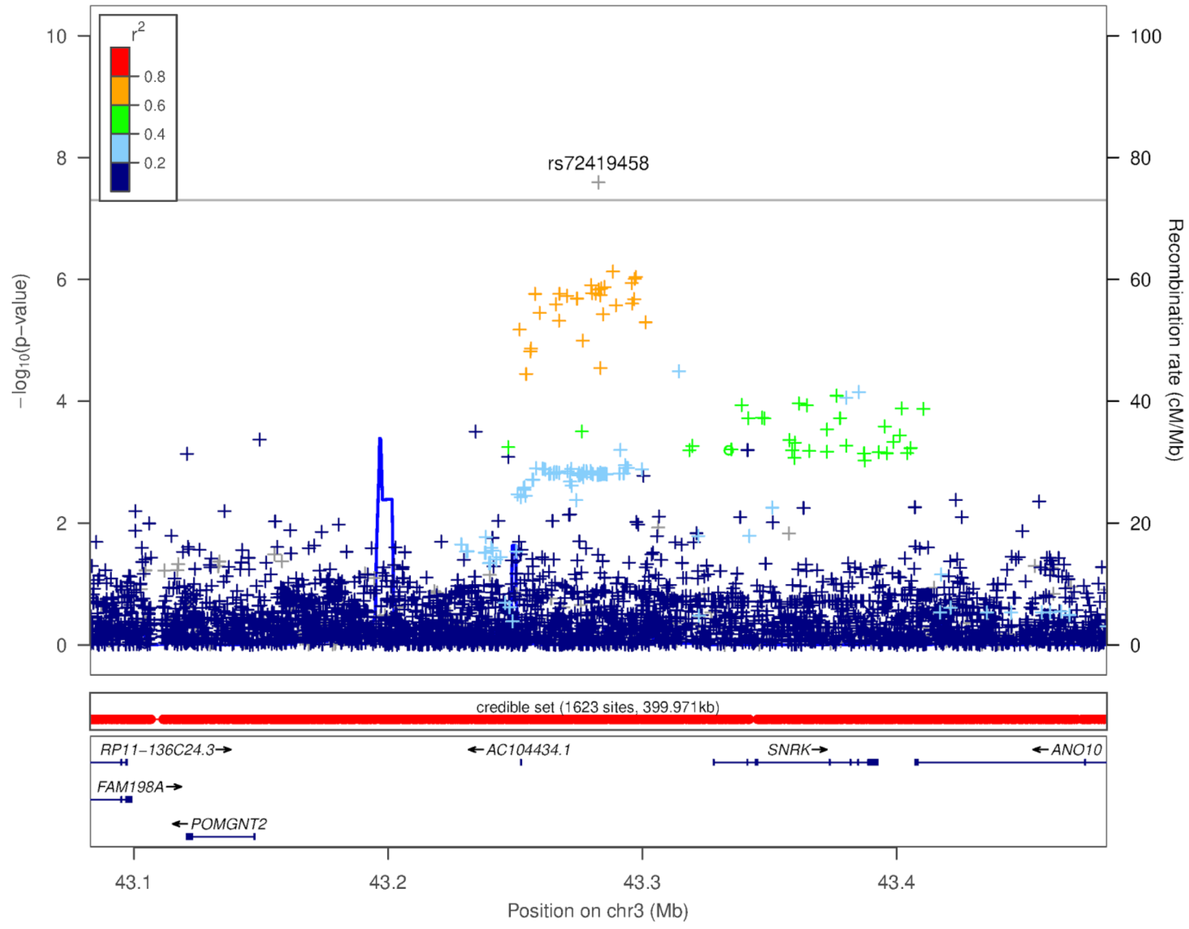


Supplementary Figure 23: Locuszoom plot for cleft lip chromosome 3 association

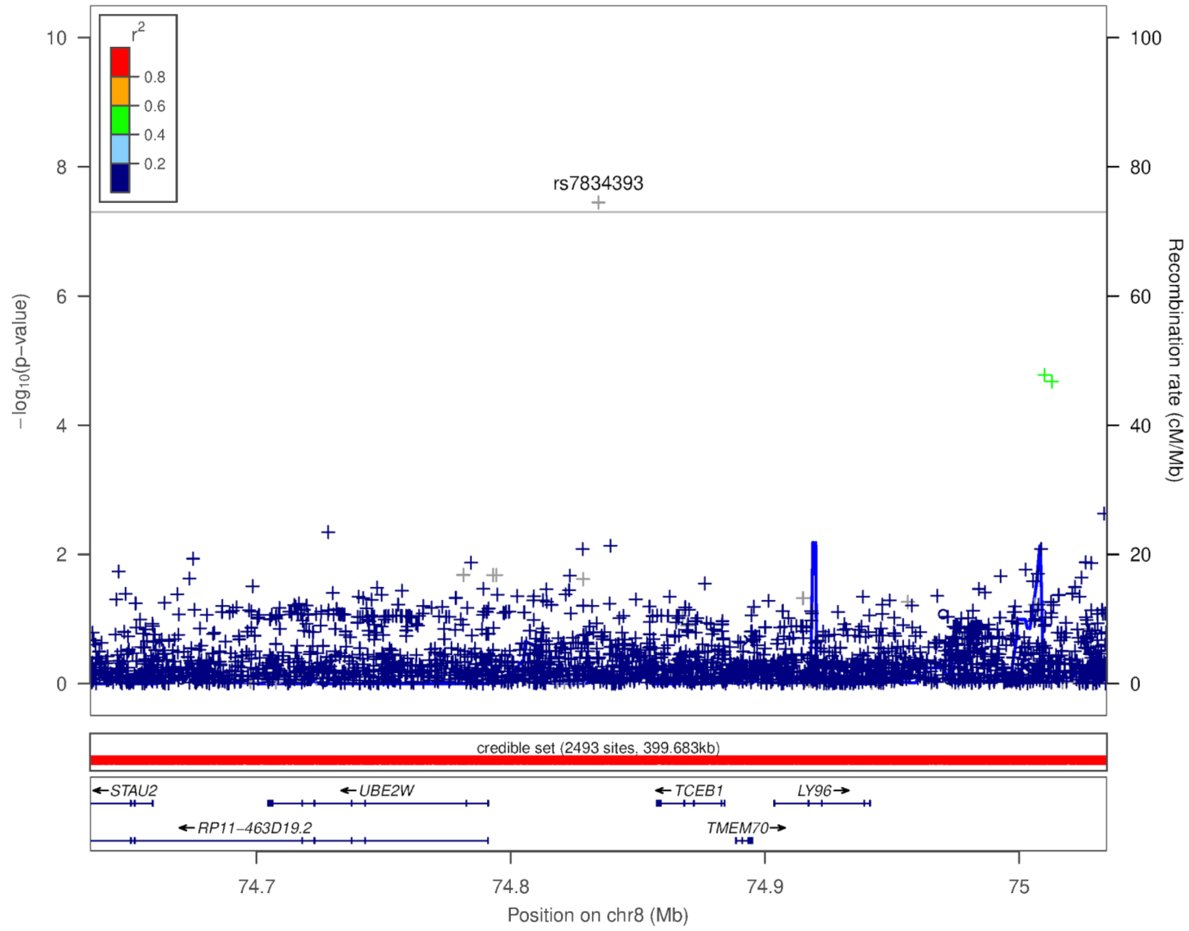


Supplementary Figure 24: Locuszoom plot for duane retraction syndrome chromosome 8 association

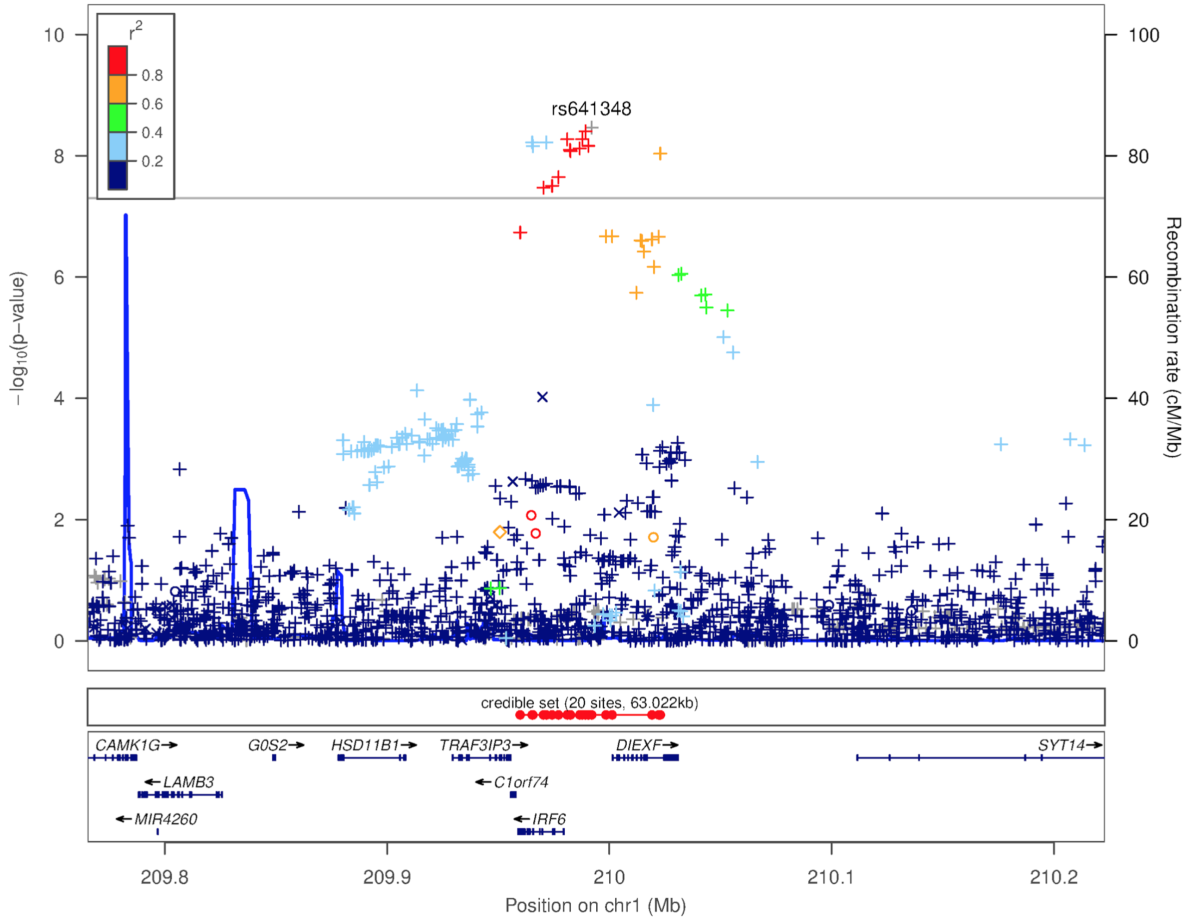


Supplementary figure 25: Locuszoom plot for cleft lip chromosome 1 association in the trans-ethnic GWAS

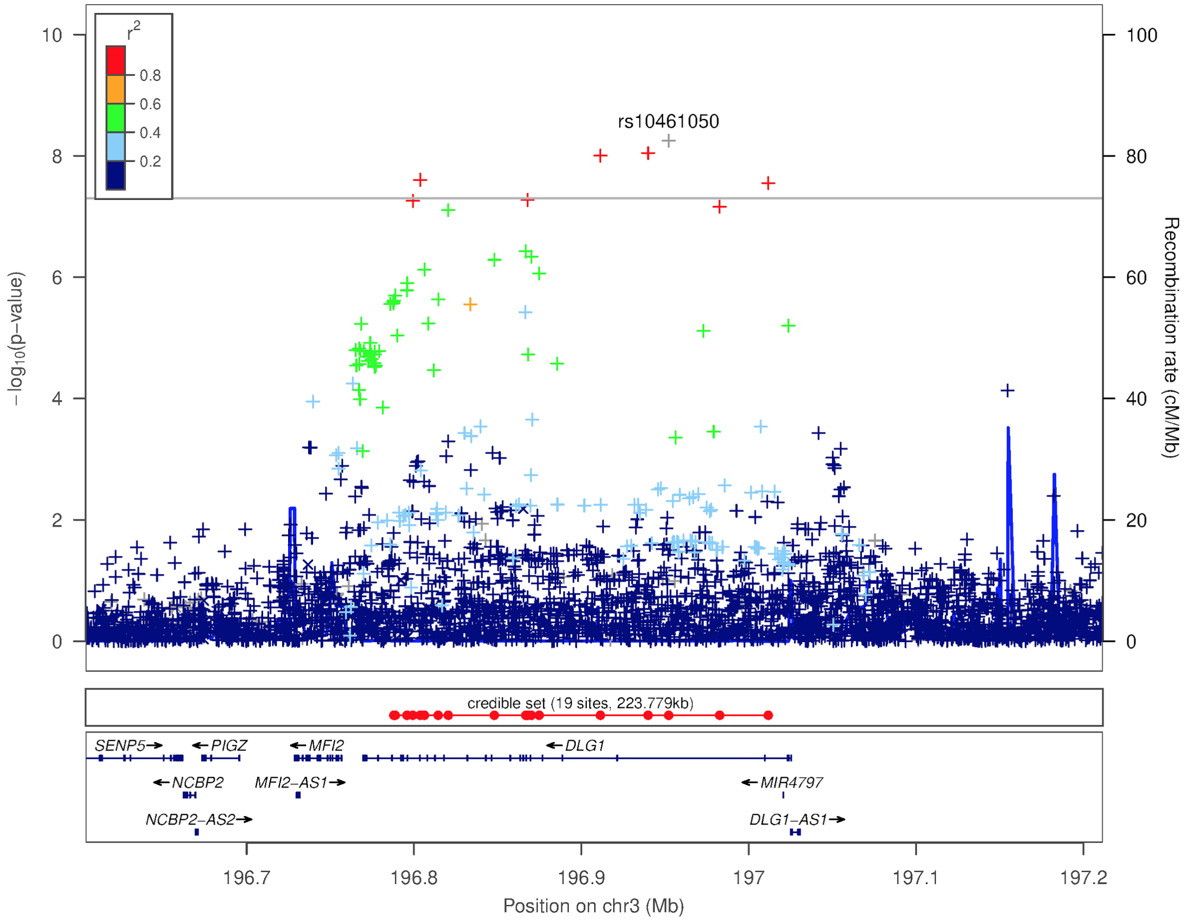


Supplementary figure 26: Locuszoom plot for cleft lip chromosome 3 association in the trans-ethnic GWAS
